# The impact of vaccination on COVID-19 outbreaks in the United States

**DOI:** 10.1101/2020.11.27.20240051

**Authors:** Seyed M. Moghadas, Thomas N. Vilches, Kevin Zhang, Chad R. Wells, Affan Shoukat, Burton H. Singer, Lauren Ancel Meyers, Kathleen M. Neuzil, Joanne M. Langley, Meagan C. Fitzpatrick, Alison P. Galvani

**Affiliations:** Agent-Based Modelling Laboratory, York University, Toronto, Ontario, M3J 1P3 Canada; Institute of Mathematics, Statistics and Scientific Computing, University of Campinas, Campinas SP, Brazil; Faculty of Medicine, University of Toronto, Toronto, Ontario, M5S 1A8 Canada; Center for Infectious Disease Modeling and Analysis (CIDMA), Yale School of Public Health, New Haven, Connecticut, USA; Emerging Pathogens Institute, University of Florida, Gainesville, FL 32610, USA; Center for Vaccine Development and Global Health, University of Maryland School of Medicine, 685 W Baltimore St, Baltimore, MD 21201 USA; Department of Integrative Biology, The University of Texas at Austin, Austin, TX 78712 USA; Canadian Center for Vaccinology, Dalhousie University, IWK Health Centre and Nova Scotia Health Authority, Halifax, Nova Scotia, B3K 6R8 Canada

**Keywords:** COVID-19, SARS-CoV-2, vaccines, outbreak simulation, United States, pandemic

## Abstract

**Background:** Global vaccine development efforts have been accelerated in response to the devastating COVID-19 pandemic. We evaluated the impact of a 2-dose COVID-19 vaccination campaign on reducing incidence, hospitalizations, and deaths in the United States (US).

**Methods:** We developed an agent-based model of SARS-CoV-2 transmission and parameterized it with US demographics and age-specific COVID-19 outcomes. Healthcare workers and high-risk individuals were prioritized for vaccination, while children under 18 years of age were not vaccinated. We considered a vaccine efficacy of 95% against disease following 2 doses administered 21 days apart achieving 40% vaccine coverage of the overall population within 284 days. We varied vaccine efficacy against infection, and specified 10% pre-existing population immunity for the base-case scenario. The model was calibrated to an effective reproduction number of 1.2, accounting for current non-pharmaceutical interventions in the US.

**Results:** Vaccination reduced the overall attack rate to 4.6% (95% CrI: 4.3% - 5.0%) from 9.0% (95% CrI: 8.4% - 9.4%) without vaccination, over 300 days. The highest relative reduction (54-62%) was observed among individuals aged 65 and older. Vaccination markedly reduced adverse outcomes, with non-ICU hospitalizations, ICU hospitalizations, and deaths decreasing by 63.5% (95% CrI: 60.3% - 66.7%), 65.6% (95% CrI: 62.2% - 68.6%), and 69.3% (95% CrI: 65.5% - 73.1%), respectively, across the same period.

**Conclusions:** Our results indicate that vaccination can have a substantial impact on mitigating COVID-19 outbreaks, even with limited protection against infection. However, continued compliance with non-pharmaceutical interventions is essential to achieve this impact.

**Key points:** Vaccination with a 95% efficacy against disease could substantially mitigate future attack rates, hospitalizations, and deaths, even if only adults are vaccinated. Non-pharmaceutical interventions remain an important part of outbreak response as vaccines are distributed over time.

## Introduction

Despite unprecedented movement restrictions, social distancing measures, and stay-at-home orders enacted in many countries [1–4], the COVID-19 pandemic has caused devastating morbidity and mortality. However, the vast majority of the global population remains susceptible to COVID-19, highlighting the need for an effective vaccine. To mitigate the mounting burden of COVID-19, vaccine development has occurred at an unprecedented pace [5]. As of December 31, 2020, safety and efficacy results for a number of vaccines have been reported [6–8], and Phase III clinical trials for several other candidates are underway [5,9–11].

Results from two large efficacy trials (Pfizer - BioNTech, Moderna) indicate a vaccine efficacy of over 90% against symptomatic and severe disease [6,7], exceeding the preferred population-based efficacy specified by the World Health Organization [12] and the United States (US) Food and Drug Administration (FDA) [13]. These vaccines have received emergency use authorization by the FDA [14,15], and vaccination has already started in the US with prioritization of healthcare workers, long-term care residents, and high-risk individuals. This compels an urgent need to understand the potential population-level impact of vaccination on COVID-19 transmission and disease outcomes.

Implementation of vaccination programs will likely take several months, depending on the ability to roll out clinics and security of vaccine supply in each state. To project the impact of vaccination and roll-out during ongoing outbreaks, we developed an age-structured transmission model, taking into account comorbidities and demographics of the US population [16–19]. We explored a strategy where healthcare workers and high-risk individuals, including those with comorbidities associated with severe COVID-19 [16,17,20] and individuals aged 65 and older, were prioritized for vaccination. This prioritization relies on the evidence that COVID-19 patients with pre-existing health conditions, including diabetes and hypertension, are 2-4 times more likely to develop severe disease than those without comorbidities [21–23]. Moreover, severity of symptoms and risk of death increase precipitously with age [24,25]. For sensitivity analyses, we varied vaccine efficacy against infection, vaccine coverage, and the level of pre-existing immunity in the population.

## Methods

### Model structure

We extended a previously developed agent-based COVID-19 transmission model to include vaccination [26]. The model encapsulates the natural history of COVID-19 with classes of individuals including: susceptible; vaccinated; latently infected (not yet infectious); asymptomatic (and infectious); pre-symptomatic (and infectious); symptomatic with either mild or severe/critical illness; recovered; and dead (Figure 1). We stratified the population into six age groups of 0-4, 5-19, 20-49, 50-64, 65-79, and 80+ years based on US demographics [19], in addition to the age-specific prevalence of comorbidities (Appendix, Table A1) [20,27]. The number of daily contacts for each individual was sampled from a negative-binomial distribution [28] with age-dependent mean and standard deviation (Appendix, Tables A2). These contacts were then distributed across age groups using an empirically-determined contact network [28].

**Figure 1.**
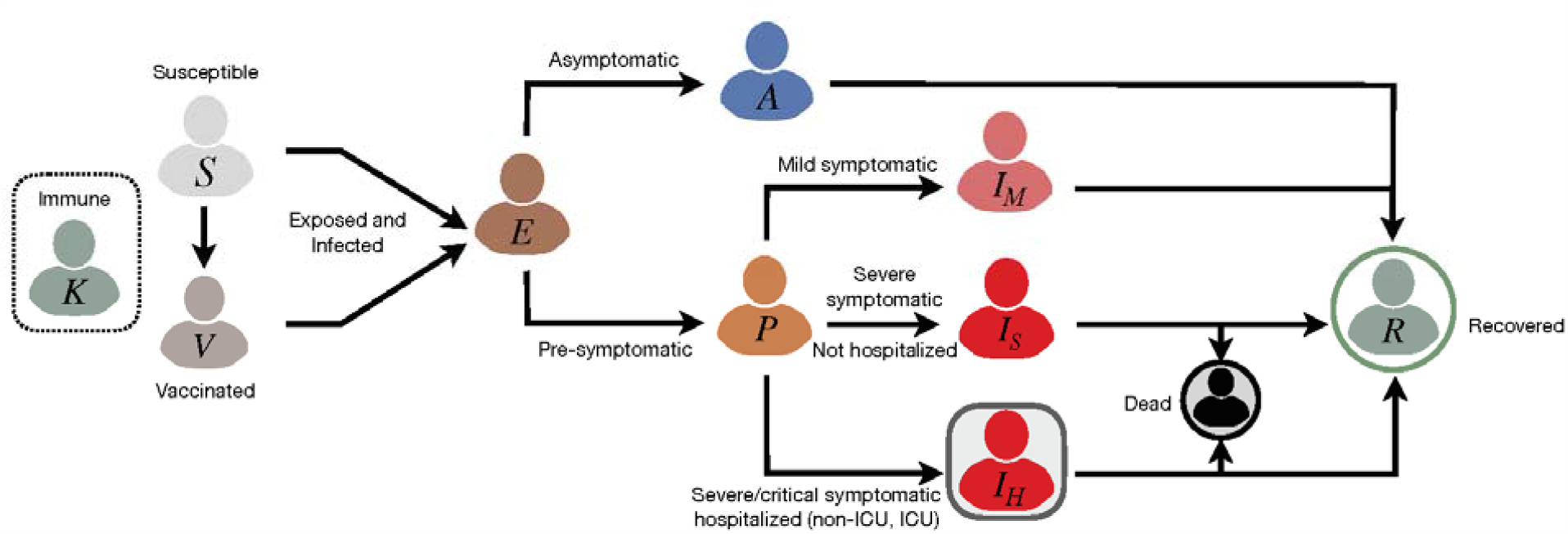
Schematic model diagram for infection dynamics and natural history of disease.

### Disease dynamics

Disease transmission was implemented probabilistically for contacts between susceptible and infectious individuals in asymptomatic, pre-symptomatic, or symptomatic stages of the disease. Based on the number of secondary cases generated during each stage of the disease [29], infectivity of mild and severe symptomatic stages was parameterized to be 44% and 89%, relative to the pre-symptomatic stage [29,30]. The infectivity of asymptomatic infection was assumed to be 26% relative to symptomatic infection, based on an average estimated 3.85 times higher incidence among close contacts of a symptomatic case compared to those of an asymptomatic individual [31]. Disease-specific parameters were sampled for each individual from their associated distributions and ranges. If infection occurred, the incubation period was sampled from a Gamma distribution with a mean of 5.2 days [32]. A proportion of infected individuals develop symptoms after a highly infectious pre-symptomatic stage [33]. The duration of the pre-symptomatic stage was sampled from a Gamma distribution with a mean of 2.3 days [30,33]. The infectious period following the onset of symptoms was sampled from a Gamma distribution with a mean of 3.2 days [34]. Symptomatic cases had an age-dependent probability of developing mild or severe/critical illness. The remaining proportion of infected individuals were asymptomatic after the latent period until recovery, with an infectious period that was sampled from a Gamma distribution with a mean of 5 days [34,35]. We assumed that recovery from a primary infection provided adequate immunity for the remainder of the simulation, preventing re-infection. A summary of model parameterization is provided in Table 1.

**Table 1.** Description of model parameters and their estimates.

| Description | 0–4 | 5–19 | 20–49 | 50–64 | 65–79 | 80+ | Source |
| --- | --- | --- | --- | --- | --- | --- | --- |
| Transmission probability per contact during pre-symptomatic stage | Depending on the level of herd immunity<br>0.0395, 0.042, 0.0465 |  |  |  |  |  | Calibrated to R=1.2 [48] |
| Incubation period (days) | LogNormal(shape: 1.434, scale: 0.661) |  |  |  |  |  | [32] |
| Asymptomatic period (days) | Gamma(shape: 5, scale: 1) |  |  |  |  |  | Derived from [34,35] |
| Pre-symptomatic period (days) | Gamma(shape: 1.058, scale: 2.174) |  |  |  |  |  | Derived from [30,33] |
| Infectious period from onset of symptoms (days) | Gamma(shape: 2.768, scale: 1.1563) |  |  |  |  |  | Derived from [34] |
| Proportion of infections that are asymptomatic | 0.30 | 0.38 | 0.33 | 0.33 | 0.19 | 0.19 | [54–56] |
| Proportion of symptomatic cases that exhibit mild symptoms | 0.95 | 0.90 | 0.85 | 0.60 | 0.20 | 0.20 | [26,37] |
| Proportion of cases hospitalized with one or more comorbidities | 37.6% |  |  |  |  |  | [16,17] |
| Non-ICU | 67% |  |  |  |  |  |  |
| ICU | 33% |  |  |  |  |  |  |
| Proportion of cases hospitalized without any comorbidities | 9% |  |  |  |  |  | [16,17] |
| Non-ICU | 75% |  |  |  |  |  |  |
| ICU | 25% |  |  |  |  |  |  |
| Length of non-ICU stay (days) | Gamma(shape: 4.5, scale: 2.75) |  |  |  |  |  | Derived from [38,39] |
| Length of ICU stay (days) | Gamma(shape: 4.5, scale: 2.75) + 2 |  |  |  |  |  | Derived from [38,39] |

### Infection outcomes

In the model, symptomatic cases with mild illness recover without the need for hospitalization, but hospital and intensive care unit (ICU) admissions were included for a proportion of severely/critically ill patients. We assumed that mild symptomatic cases and severely ill individuals who were not hospitalized practice self-isolation immediately upon symptom onset. The contact patterns during isolation were specified by an age-dependent daily number of contacts based on a matrix derived from a representative sample population during COVID-19 lockdown [36]. Non-ICU and ICU admissions of patients were parameterized based on age-stratified COVID-19 hospitalization data, and the presence of comorbidities [16,17]. For those who were hospitalized, time from symptom onset to admission was sampled in the range of 2-5 days [26,37]. The lengths of non-ICU and ICU stays were sampled from Gamma distributions with means of 12.4 and 14.4 days, respectively [38,39].

### Vaccination

We implemented a two-dose vaccination campaign achieving 40% coverage of the entire population within 284 days. We assumed that 70% was the maximum achievable coverage in any age group, with an age-dependent distribution similar to seasonal influenza vaccination in the US [40]. Vaccines were prioritized to the following groups sequentially: (i) healthcare workers (5% of the total population [41]), adults with comorbidities, and those aged 65 and older (i.e., protection cohort); and (ii) all other individuals aged 18-64 (i.e., disruption minimization cohort) [42]. Comorbidities included cardiovascular disease, diabetes, asthma, chronic obstructive pulmonary disease, hypertension, and cancer [20]. Pre-existing immunity or contemporaneous infection with COVID-19 was not a factor in vaccine allocation. The age-specific coverage resulting from this prioritization was 48% of those aged 18-49, 48% of those aged 50-64, and 70% of those aged 65+. We specified a roll-out strategy in which 30 individuals per 10,000 population would be vaccinated per day, corresponding to 6.93 million vaccine doses per week for the entire US population, for approximately 41 weeks. Vaccination occurred during this time period to reach 40% coverage and outcomes were evaluated for 300 days. Infection dynamics continued during the simulations for susceptible and vaccinated individuals.

We included a 21-day interval between the first and second vaccine doses [6]. The vaccine efficacy (*V*_*e*_) against symptomatic and severe disease was assumed to be 52%, 14 days after the first dose, and 95%, 1 week after the second dose [6]. In the absence of data for vaccine efficacy against infection or transmission, we assumed that the vaccine protection against infection was 50% lower than its efficacy against disease (base-case), with additional scenarios of (i) 0%; and (ii) the same efficacy against disease, after each dose of the vaccine. We further simulated the model for these scenarios with a 28-day interval between the two doses [43]. Vaccine efficacy against infection was implemented as a reduction in the probability of transmission when a vaccinated individual encountered an infectious individual. This efficacy was reduced by a factor of *q* in vaccinated individuals with any comorbidities or in persons older than 65 years of age, where *q* was sampled uniformly from the 10-50% range for each individual. This parameterization was based on observed reductions in influenza vaccine effectiveness among frail and comorbid individuals [44,45]. For these individuals, we also assumed that vaccine efficacy against disease was reduced by the same factor *q*, if infection occurred post-vaccination, thereby affecting hospitalization and death rates. As a sensitivity analysis, we considered vaccination scenarios without reduction of vaccine efficacy in these individuals. The immunity conferred by vaccination or infection was assumed to last longer than one year (i.e., beyond the simulation timelines).

### Model scenarios

In the base-case scenario, we assumed a 10% level of pre-existing immunity in the population at the onset of simulations, within the range of estimates provided in recent seroprevalence studies [46,47]. In scenario analysis, 5% pre-existing immunity was considered to represent regions that have not yet been substantially affected by COVID-19 outbreaks, and alternatively 20% pre-existing immunity was used to represent the expectation that immunity will continue to accrue prior to vaccine availability. To accurately capture the age distribution of population immunity, the model was simulated in the absence of vaccination in an entirely susceptible population. Then, the infection rates in different age groups were derived when the overall attack rate reached 5%, 10%, and 20%, and the corresponding distributions were used as the starting population for the vaccination model (Appendix, Table A3). Additional scenarios for vaccination coverages in the range 20-60% are also presented in the Appendix.

### Model implementation

Model calibration was performed using an effective reproduction number of 1.2 to account for the effect of current non-pharmaceutical COVID-19 interventions in the US [48]. Simulations were seeded with three initial cases in the pre-symptomatic stage in a population of 10,000 individuals (a scalable size in agent-based modelling) for different levels of herd immunity, and the results averaged over 1000 independent Monte-Carlo realizations, which was sufficient for stabilization. Credible intervals (CrI) were obtained using the bias-corrected and accelerated bootstrap method. The model was implemented in Julia language and is available at: https://github.com/thomasvilches/covid_vac.

## Results

The transmission probability per contact was calibrated to an effective reproduction number *R*_e_=1.2 [48]. For the base-case scenario of 10% pre-existing immunity, and with self-isolation of infected individuals following symptom onset, the attack rate was projected to be 9.0% (95% CrI: 8.4% - 9.5%) on day 300 in the absence of a vaccine.

### Attack rate

Vaccination with 10% pre-existing immunity, even with a 10-50% reduction of vaccine efficacy in elderly and comorbid individuals, substantially mitigated the attack rate across all age groups (Figure 2A), with a mean overall attack rate of 4.6% (95% CrI: 4.3% - 5.0%) on day 300 (Figure 3). Achieving approximately 50% reduction compared to the no vaccination scenario, the vaccination program would avert 435 (95% CrI: 371 - 494) infections per 10,000 people over 300 days from the start of vaccine distribution. The attack rate was most substantially reduced among individuals aged 65+, by 54-62% (Figure 2). Although no children under 18 years of age were vaccinated in this model, the attack rate among those under 20 years of age was reduced by at least 36%, largely driven by indirect protection and reduced incidence among adults. Sensitivity analyses for attack rates corresponding to 5% and 20% pre-existing immunity also revealed significant decreases attributed to vaccination across all age groups, but the impact of vaccination was reduced at higher levels of pre-existing immunity (Figure 2).

**Figure 2.**
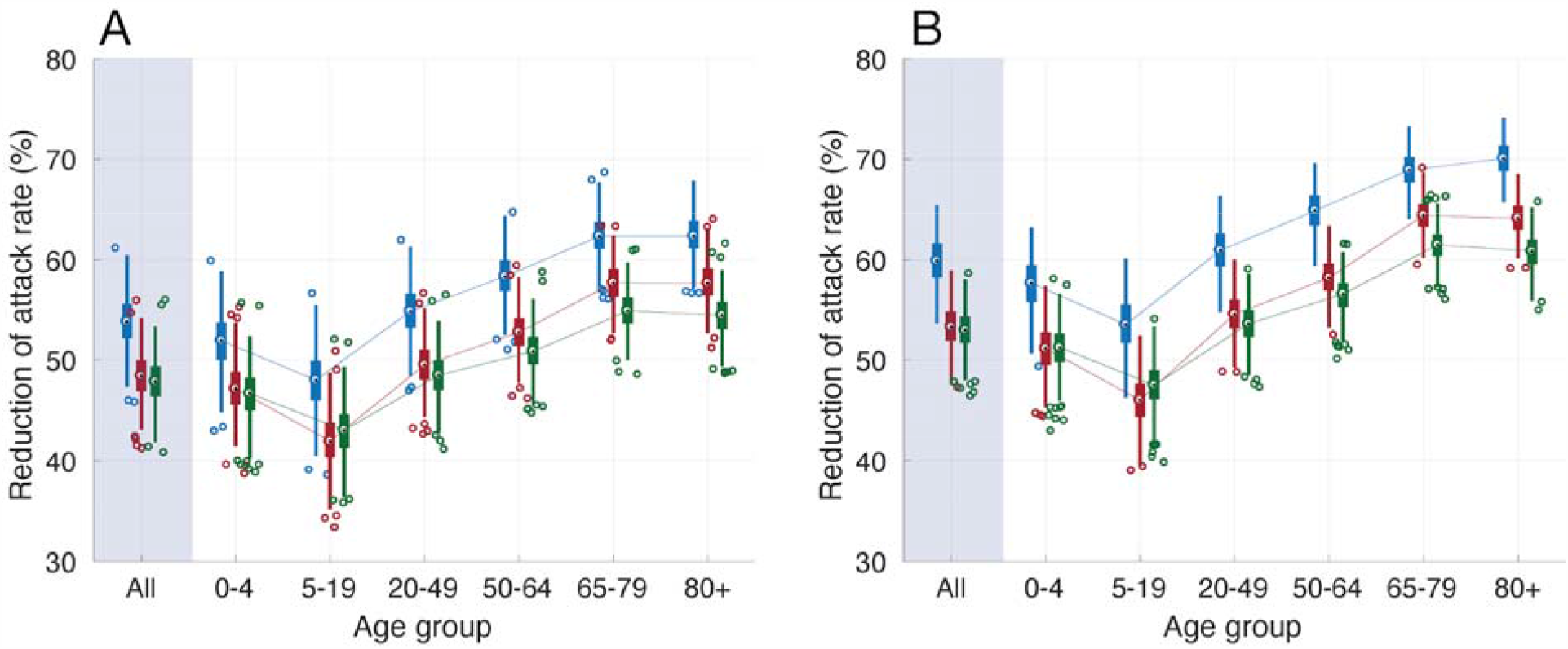
Overall and age-specific relative reduction of mean attack rates with vaccination, as compared to the outbreak scenario in the absence of vaccination, with 5% (blue), 10% (red), and 20% (green) levels of pre-existing immunity over 300 days. Panels (A) and (B) correspond, respectively, to scenarios with and without reduction of vaccine efficacy in comorbid individuals and the elderly.

**Figure 3.**
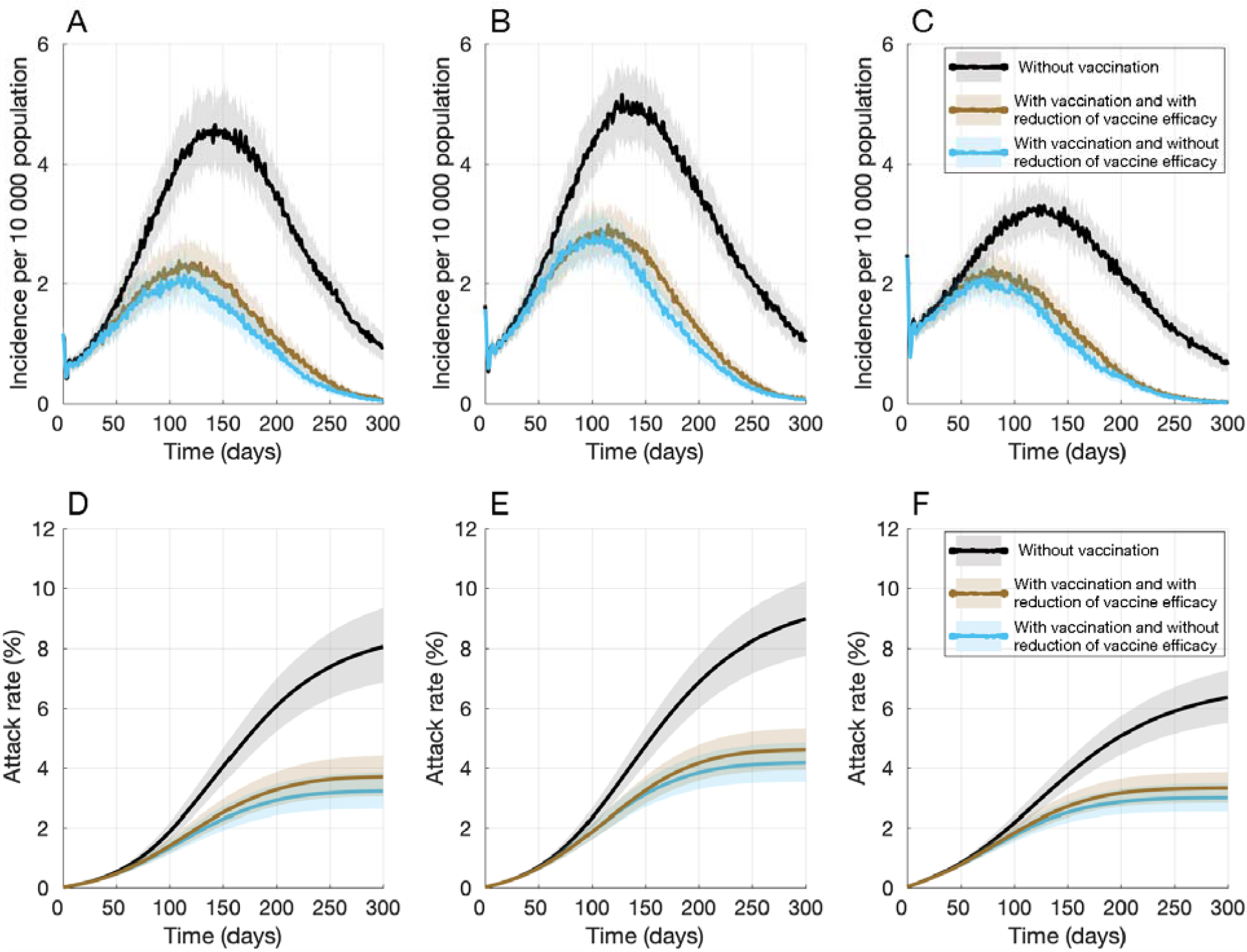
Projected daily incidence of COVID-19 per 10,000 population with 5% (A), 10% (B), and 20% (C) levels of pre-existing immunity. Projected temporal attack rates with 5% (D), 10% (E), and 20% (F) levels of pre-existing immunity over 300 days. Vaccination started on day 0. Colored curves with vaccination correspond, respectively, to scenarios with (brown) and without (blue) reduction of vaccine efficacy in comorbid individuals and the elderly.

When vaccine efficacy in elderly and comorbid individuals was the same as in other subpopulations, vaccination led to a higher decrease in attack rates across all age groups (Figure 2). In this scenario, the comparative advantage of vaccination in reducing incidence and the overall attack rate was diminished as the level of pre-existing immunity in the population increased (Figure 3).

We observed that, in the absence of vaccination, the daily incidence remained above 1 per 10,000 population for at least 288 days (Figure 3). However, vaccination with 40% coverage reduced the outbreak peak and led to a daily incidence below 1 case 2-3 months earlier, within 203 - 222 days from the start of vaccination. This earlier control of the outbreak in the base-case scenario with 10% pre-existing immunity was also observed at other levels of pre-existing immunity (Figure 3).

### Hospitalizations and Deaths

In the absence of vaccination, and with 10% pre-existing immunity, total non-ICU and ICU hospitalizations were projected to be 20.3 (95% CrI: 19.0 - 21.4) and 9.3 (95% CrI: 8.7 - 9.9) per 10,000 population, respectively, with 2.3 (95% CrI: 2.1 - 2.4) deaths per 10,000 population on day 300 (Figure 4).

**Figure 4.**
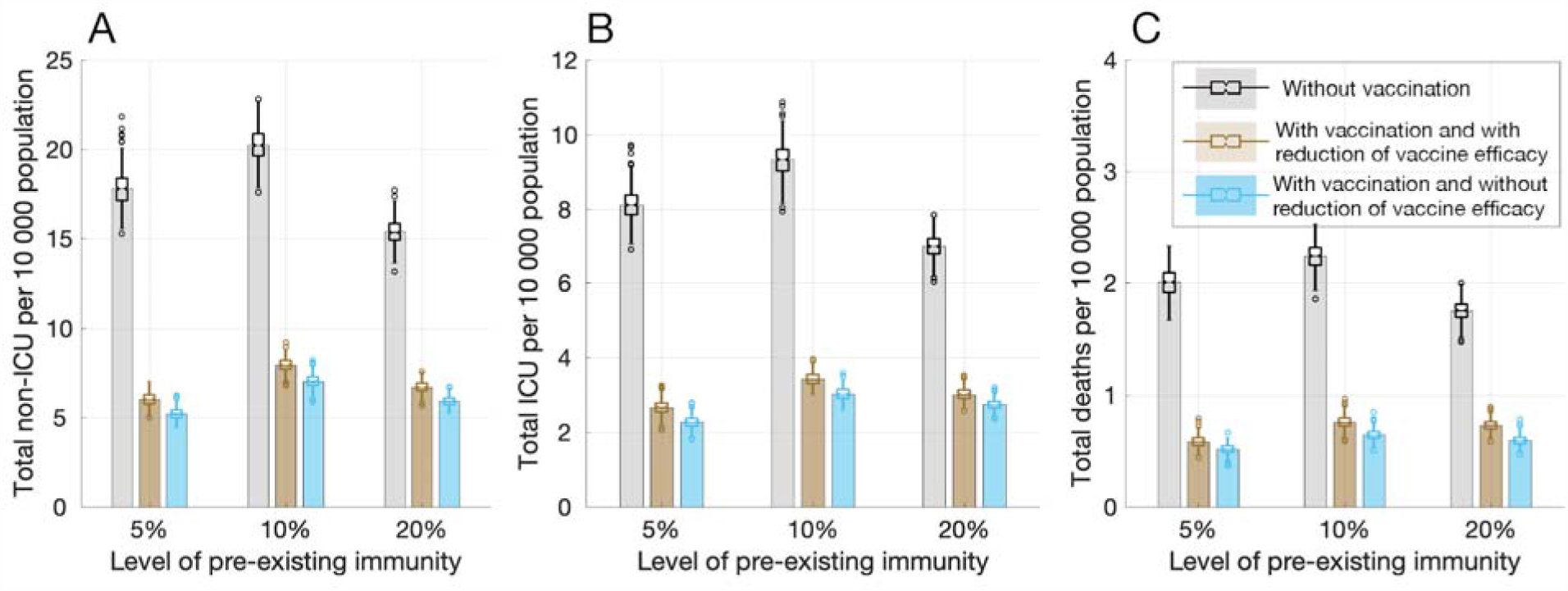
Projected total number of non-ICU hospitalizations (A), ICU hospitalizations (B), and deaths (C) per 10,000 populations with 5%, 10%, and 20% levels of pre-existing immunity over 300 days. Colored bars with vaccination correspond, respectively, to scenarios with (brown) and without (blue) reduction of vaccine efficacy in comorbid individuals and the elderly.

Vaccination with reduced efficacy in elderly and comorbid individuals still markedly reduced hospitalizations and deaths (Figure 4). Non-ICU hospitalizations, ICU hospitalizations, and deaths would be reduced by 63.5% (95% CrI: 60.3% - 66.7%), 65.6% (95% CrI: 62.2% - 68.6%), and 69.3% (95% CrI: 65.5% - 73.1%), respectively, over 300 days from the start of vaccination. We projected that vaccination would lead to higher reductions in hospitalizations and deaths for 5% pre-existing immunity. However, vaccine attributable reductions in non-ICU hospitalizations, ICU hospitalizations, and deaths were lower with 20% pre-existing immunity, and were projected to be 59.5% (95% CrI: 55.8% - 62.9%), 59.5% (95% CrI: 55.6% - 63.0%), and 61.6% (95% CrI: 57.0% - 66.2%), respectively (Figure 4). If vaccine efficacy was not reduced in elderly and comorbid individuals, the effect of vaccination in reducing hospitalizations and deaths was increased. However, similar to the case of attack rates, this additional benefit was diminished with higher levels of pre-existing immunity (Figure 4).

We performed sensitivity analyses with additional scenarios corresponding to vaccine coverages in the range 10-60%, different protection efficacy of vaccine against infection, and when the time interval between the two vaccine doses was 28 days. Results of these scenarios, summarized in Appendix, present qualitatively similar outcomes for different levels of population immunity (Appendix, Sections 4 and 5). There was no substantial effect of the one week difference in dosing, but a greater reduction of disease burden was achieved with higher vaccine efficacy against infection.

## Discussion

COVID-19 outbreaks have caused significant global morbidity and mortality, in addition to undermining the economic and social well-being of individuals and communities. Despite this devastating toll, the majority of the population remains susceptible to SARS-CoV-2 infection [49]. Thus, vaccine development has been a high priority. The scale and speed of vaccine development efforts have been unprecedented, and highly protective vaccines are beginning to be distributed. This study shows that COVID-19 vaccines with 95% efficacy in preventing disease, even if they conferred limited protection against infection, could substantially mitigate future attack rates, hospitalizations, and deaths.

Our findings should be interpreted within study assumptions and limitations. First, our model vaccinated a large proportion of high-risk individuals, including 70% of healthcare workers [50] and 56% of comorbid individuals. Although this coverage may be difficult to achieve in the short term [51], strategic public health campaigns and transparent communication regarding vaccine safety may be able to improve uptake. Second, we assumed that all vaccinated individuals were willing to receive both doses. If substantial drop-out occurs after the first dose, vaccines could be used more quickly for the general population, and the short-term effect of drop-outs may be minor. Third, the daily number of contacts in the model was age-dependent without consideration of the location of occurrence (e.g., within households, workplaces, and schools). However, this is not expected to change our community-based results, since model calibration would modulate the transmission probability for a given reproduction number. Similarly, any reduction of contacts and variation in contact patterns are accounted for in the calibration process when transmission probability is determined. Finally, the model did not explicitly simulate other mitigation measures (e.g., social distancing, mask-wearing, testing, and contact tracing); however, we calibrated the model to current estimates of the effective reproduction number to account for known compliance with such measures in the US. Further studies are needed to determine the vaccine coverage required to eliminate the need for non-pharmaceutical interventions.

Given the limited population-level immunity to COVID-19 [46], vaccination remains a key preventive measure to reduce disease burden and mitigate future outbreaks. Our study suggests that a vaccine could have a substantial impact on reducing incidence, hospitalizations, and deaths, especially among vulnerable individuals with comorbidities and risk factors associated with severe COVID-19. Thus, mobilizing public health resources is imperative to achieve the proposed goal of distributing 100 million vaccine doses over 100 days in the US population by the incoming administration [52]. Our findings support the Advisory Committee on Immunization Practices recommendations [53], highlighting that a targeted vaccination strategy can effectively mitigate disease burden and the societal impact of COVID-19. We also find that, even with the relatively rapid roll-out simulated here, it may take several months to control COVID-19 at the population level. Moreover, this impact is achieved in the context of continued public health efforts, and is not possible without diligent attention to the other aspects of infection prevention and control such as masking, hand hygiene, testing, contact-tracing, and isolation of infected cases. If current vaccination programs are accompanied by widespread relaxation of other measures, a much higher coverage will be necessary with a significantly higher distribution capacity. Nevertheless, our results are an encouraging signal of the power and promise of vaccines against COVID-19.

## Data Availability

The model was implemented in Julia language and is available at: https://github.com/thomasvilches/covid_vac.

## Reproducibility Statement

The computational system and parameters are available under an open-source license at: https://github.com/thomasvilches/covid_vac.

