## Supplementary File for "The impact of vaccination on COVID-19 outbreaks in the United States"

This appendix provides further details of model parameterization, projections of attack rates and outcomes with vaccination, as well as sensitivity analyses with varying parameters of vaccine coverage, vaccine efficacy against infection, time interval between the first and second vaccine doses, and results of simulations with and without a reduction of vaccine efficacy in elderly and comorbid individuals.

### 1. Vaccination

Vaccination campaigns started with distribution of vaccines to healthcare workers (5% of the total population), individuals with comorbidities older than 18 years of age (Table A1), and those aged 65 and older. Healthcare workers included persons employed in ambulatory care, hospitals, nursing homes, and long-term care facilities (1). Comorbidities considered in vaccine prioritization included cardiovascular disease, diabetes, asthma, chronic obstructive pulmonary disease, hypertension, and cancer (2). On each day, 30 doses of vaccine per 10,000 population were distributed, with a second dose being offered after the specified time interval (either 21 or 28 days after the first dose). Second priority groups included individuals aged 18-64, and vaccines were distributed randomly to reach the overall coverage of 40% for both vaccine doses (or the specified coverage in the sensitivity analyses).

### 2. Model parameters

**Table A1.** Percentage of population in different age groups with and without comorbidities.

| Condition | 0-4 | 5-19 | 20-49 | 50-64 | 65+ | All age groups |
|---|---|---|---|---|---|---|
| With comorbidity | 5% | 10% | 28% | 55% | 78% | 37% |
| Without comorbidity | 95% | 90% | 72% | 45% | 22% | 63% |
| **Fraction of total population** | 5.9% | 18.9% | 39.6% | 18.9% | 16.6% | 100% |

The number of daily contacts for each agent was sampled from an age-specific negative binomial distribution and contacts were distributed among different age groups based on a contact matrix (Table A2) (3). For reduced contacts due to interventions (i.e., self-isolation), we used estimates of contacts derived from a recent study quantifying the impact of physical distancing during the COVID-19 lockdown in the UK (4). Measures of lockdown included school closure, quarantine and isolation of cases, and physical distancing in workplaces. The reduction of daily contacts for self-isolated individuals was at least 70% in different age groups. The overall effect of contact patterns was implicitly accounted for in the model calibration to the effective reproduction number.

**Table A2.** Mixing patterns and the daily number of contacts derived from empirical observations. Daily numbers of contacts were sampled from negative binomial distributions for different scenarios.

| Age group | Proportion of contacts between age groups | | | | | Number of daily contacts without self-isolation Mean (SD) Ref: (3) | Number of daily contacts for self-isolated individuals Mean (SD) Ref: (4) |
|---|---|---|---|---|---|---|---|
| | 0-4 | 5-19 | 20-49 | 50-65 | 65+ | | |
| **0-4** | 0.2287 | 0.1839 | 0.4219 | 0.1116 | 0.0539 | 10.21 (7.65) | 2.86 (2.14) |

| | | | | | | | |
|---|---|---|---|---|---|---|---|
| **5-19** | 0.0276 | 0.5964 | 0.2878 | 0.0591 | 0.0291 | 16.793 (11.7201) | 4.70 (3.28) |
| **20-49** | 0.0376 | 0.1454 | 0.6253 | 0.1423 | 0.0494 | 13.795 (10.5045) | 3.86 (2.95) |
| **50-65** | 0.0242 | 0.1094 | 0.4867 | 0.2723 | 0.1074 | 11.2669 (9.5935) | 3.15 (2.66) |
| **65+** | 0.0207 | 0.1083 | 0.4071 | 0.2193 | 0.2446 | 8.0027 (6.9638) | 2.24 (1.95) |

**Table A3.** Distribution of the levels of pre-existing immunity in different age groups before vaccine implementation.

| **Pre-existing immunity** | **0-4** | **5-19** | **20-49** | **50-64** | **65-79** | **80+** |
|---|---|---|---|---|---|---|
| 5% | 1.5% | 7.9% | 6.6% | 3.6% | 0.5% | 0.9% |
| 10% | 5.4% | 14.8% | 12.4% | 7.5% | 3.3% | 3.5% |
| 20% | 11.9% | 28.2% | 24.3% | 16.0% | 8.0% | 8.1% |

**Table A4.** Vaccination coverage of different age groups.

| **Age group** | **0-17** | **18-49** | **50-64** | **65-79** | **80+** |
|---|---|---|---|---|---|
| Vaccination coverage | 0% | 48% | 48% | 70% | 70% |

### 3. Estimates of attack rates, hospitalizations, and deaths

Estimates in this section correspond to the results presented in the main text with 40% vaccination coverage, and a 21-day time interval between the two vaccine doses. Vaccine efficacy against infection was 50% lower than the efficacy against disease after each dose, and this efficacy was reduced by 10-50% in vaccinated elderly and comorbid individuals.

**Table A5.** Projected age-specific and overall attack rates per 10,000 population without vaccination and with 40% vaccination coverage over 300 days, corresponding to Figures 2 and 3 of the main text.

| Age group | 0-4 | 5-19 | 20-49 | 50-64 | 65-79 | 80+ | All |
|---|---|---|---|---|---|---|---|
| **Pre-existing immunity** | **Mean projected attack rate % (95% CrI) without vaccination** | | | | | | |
| 5% | 4.1<br>3.8 - 4.4 | 11.7<br>10.9 - 12.7 | 10.0<br>9.3 - 10.7 | 6.1<br>5.7 - 6.6 | 2.8<br>2.6 - 3.0 | 2.8<br>2.6 - 3.1 | 8.0<br>7.4 - 8.6 |
| 10% | 4.7<br>4.4 - 5.0 | 12.6<br>11.8 - 13.3 | 11.2<br>10.5 - 11.8 | 7.0<br>6.6 - 7.4 | 3.3<br>3.1 - 3.5 | 3.3<br>3.1 - 3.5 | 9.0<br>8.4 - 9.5 |
| 20% | 3.5<br>3.3 - 3.7 | 8.5<br>8.0 - 8.9 | 7.9<br>7.5 - 8.3 | 5.3<br>4.9 - 5.6 | 2.6<br>2.4 - 2.8 | 2.6<br>2.4 - 2.7 | 6.4<br>6.0 - 6.7 |
| | **Mean projected attack rate % (95% CrI) with vaccination** | | | | | | |
| 5% | 2.0<br>1.8 - 2.2 | 6.1<br>5.6 - 6.5 | 4.5<br>4.1 - 4.9 | 2.5<br>2.3 - 2.8 | 1.1<br>1.0 - 1.2 | 1.1<br>1.0 - 1.2 | 3.7<br>3.4 - 4.0 |
| 10% | 2.4<br>2.3 - 2.6 | 7.3<br>6.8 - 7.9 | 5.6<br>5.2 - 6.0 | 3.3<br>3.1 - 3.5 | 1.4<br>1.3 - 1.5 | 1.4<br>1.3 - 1.5 | 4.6<br>4.3 - 5.0 |
| 20% | 1.9<br>1.7 - 2.0 | 4.8<br>4.5 - 5.2 | 4.1<br>3.8 - 4.3 | 2.6<br>2.4 - 2.7 | 1.2<br>1.1 - 1.2 | 1.2<br>1.1 - 1.3 | 3.3<br>3.1 - 3.5 |

**Table A6.** Projected age-specific and overall non-ICU hospitalizations per 10,000 population without vaccination and with 40% vaccination coverage over 300 days, corresponding to Figure 4 of the main text.

| Age group | 0-4 | 5-19 | 20-49 | 50-64 | 65-79 | 80+ | All |
|---|---|---|---|---|---|---|---|
| **Pre-existing immunity** | **Mean projected total non-ICU hospitalizations (95% CrI) without vaccination** | | | | | | |
| 5% | 0.1<br>0.1 - 0.1 | 1.2<br>1.1 - 1.3 | 4.7<br>4.3 - 5.0 | 5.4<br>5.0 - 5.8 | 2.6<br>2.4 - 2.9 | 3.8<br>3.5 - 4.2 | 17.8<br>16.5 - 19.1 |
| 10% | 0.1<br>0.1 - 0.1 | 1.3<br>1.2 - 1.4 | 5.3<br>4.9 - 5.6 | 6.3<br>5.9 - 6.7 | 3.0<br>2.8 - 3.3 | 4.4<br>4.1 - 4.7 | 20.3<br>18.9 - 21.6 |
| 20% | 0.1<br>0.0 - 0.1 | 0.8<br>0.8 - 0.9 | 3.7<br>3.5 - 3.9 | 4.9<br>4.6 - 5.3 | 2.4<br>2.3 - 2.6 | 3.4<br>3.2 - 3.6 | 15.3<br>14.4 - 16.3 |

| | | | | | | | |
|---|---|---|---|---|---|---|---|
| | Mean projected total non-ICU hospitalizations (95% CrI) with vaccination | | | | | | |
| 5% | 0.0<br>0.0 - 0.1 | 0.6<br>0.5 - 0.7 | 1.8<br>1.6 - 1.9 | 1.9<br>1.7 - 2.1 | 0.7<br>0.6 - 0.8 | 1.0<br>0.9 - 1.1 | 6.0<br>5.5 - 6.5 |
| 10% | 0.0<br>0.0 - 0.0 | 0.7<br>0.6 - 0.8 | 2.2<br>2.1 - 2.4 | 2.5<br>2.3 - 2.7 | 1.0<br>0.9 - 1.1 | 1.4<br>1.3 - 1.5 | 7.9<br>7.3 - 8.5 |
| 20% | 0.0<br>0.0 - 0.0 | 0.5<br>0.4 - 0.6 | 1.7<br>1.5 - 1.8 | 2.3<br>2.2 - 2.5 | 0.9<br>0.8 - 1.0 | 1.3<br>1.2 - 1.4 | 6.6<br>6.2 - 7.1 |

**Table A7.** Projected age-specific and overall ICU hospitalizations per 10,000 population without vaccination and with 40% vaccination coverage over 300 days, corresponding to Figure 4 of the main text.

| Age group | 0-4 | 5-19 | 20-49 | 50-64 | 65-79 | 80+ | All |
|---|---|---|---|---|---|---|---|
| Pre-existing immunity | Mean projected total ICU hospitalizations (95% CrI) without vaccination | | | | | | |
| 5% | 0.0<br>0.0 - 0.0 | 0.5<br>0.4 - 0.5 | 2.1<br>2.0 - 2.3 | 2.5<br>2.3 - 2.7 | 1.3<br>1.2 - 1.5 | 1.7<br>1.6 - 1.9 | 8.1<br>7.5 - 8.7 |
| 10% | 0.0<br>0.0 - 0.0 | 0.5<br>0.4 - 0.5 | 2.2<br>2.0 - 2.3 | 3.0<br>2.8 - 3.2 | 1.5<br>1.4 - 1.6 | 2.2<br>2.0 - 2.4 | 9.3<br>8.7 - 10.0 |
| 20% | 0.0<br>0.0 - 0.0 | 0.3<br>0.3 - 0.3 | 1.6<br>1.5 - 1.7 | 2.3<br>2.1 - 2.5 | 1.2<br>1.1 - 1.3 | 1.6<br>1.5 - 1.7 | 7.0<br>6.5 - 7.4 |
| | Mean projected total ICU hospitalizations (95% CrI) with vaccination | | | | | | |
| 5% | 0.0<br>0.0 - 0.0 | 0.2<br>0.2 - 0.2 | 0.8<br>0.7 - 0.8 | 0.8<br>0.8 - 0.9 | 0.3<br>0.3 - 0.4 | 0.5<br>0.4 - 0.6 | 2.7<br>2.4 - 3.0 |
| 10% | 0.0<br>0.0 - 0.0 | 0.3<br>0.3 - 0.3 | 0.9<br>0.9 - 1.0 | 1.1<br>1.0 - 1.2 | 0.5<br>0.4 - 0.5 | 0.6<br>0.6 - 0.7 | 3.4<br>3.1 - 3.7 |
| 20% | 0.0<br>0.0 - 0.0 | 0.2<br>0.2 - 0.2 | 0.7<br>0.6 - 0.8 | 1.1<br>1.0 - 1.2 | 0.5<br>0.4 - 0.5 | 0.6<br>0.5 - 0.7 | 3.0<br>2.8 - 3.2 |

**Table A8.** Projected age-specific and overall deaths per 10,000 population without vaccination and with 40% vaccination coverage over 300 days, corresponding to Figure 4 of the main text.

| Age group | 0-4 | 5-19 | 20-49 | 50-64 | 65-79 | 80+ | All |
|---|---|---|---|---|---|---|---|
| Pre-existing immunity | Mean projected total deaths (95% CrI) without vaccination | | | | | | |

| | | | | | | | |
|---|---|---|---|---|---|---|---|
| 5% | 0.0<br>0.0 - 0.0 | 0.0<br>0.0 - 0.0 | 0.2<br>0.1 - 0.2 | 0.3<br>0.3 - 0.3 | 0.4<br>0.3 - 0.4 | 1.1<br>1.0 - 1.2 | 2.0<br>1.8 - 2.2 |
| 10% | 0.0<br>0.0 - 0.0 | 0.0<br>0.0 - 0.0 | 0.2<br>0.1 - 0.2 | 0.4<br>0.4 - 0.4 | 0.4<br>0.3 - 0.4 | 1.3<br>1.2 - 1.4 | 2.2<br>2.1 - 2.4 |
| 20% | 0.0<br>0.0 - 0.0 | 0.0<br>0.0 - 0.0 | 0.1<br>0.1 - 0.2 | 0.3<br>0.2 - 0.3 | 0.4<br>0.3 - 0.4 | 1.0<br>0.9 - 1.0 | 1.8<br>1.6 - 1.9 |
| | **Mean projected total deaths (95% CrI) with vaccination** | | | | | | |
| 5% | 0.0<br>0.0 - 0.0 | 0.0<br>0.0 - 0.0 | 0.1<br>0.1 - 0.1 | 0.1<br>0.1 - 0.2 | 0.1<br>0.1 - 0.1 | 0.3<br>0.2 - 0.3 | 0.6<br>0.5 - 0.7 |
| 10% | 0.0<br>0.0 - 0.0 | 0.0<br>0.0 - 0.0 | 0.1<br>0.1 - 0.1 | 0.2<br>0.1 - 0.2 | 0.1<br>0.1 - 0.2 | 0.4<br>0.3 - 0.4 | 0.8<br>0.7 - 0.8 |
| 20% | 0.0<br>0.0 - 0.0 | 0.0<br>0.0 - 0.0 | 0.1<br>0.1 - 0.1 | 0.1<br>0.1 - 0.2 | 0.1<br>0.1 - 0.2 | 0.4<br>0.3 - 0.4 | 0.7<br>0.7 - 0.8 |

### 4. Sensitivity analyses for vaccination scenarios with different coverages and protection efficacies against infection. Time interval between the two vaccine doses is 21 days.

**Part 1:** Vaccine efficacy was reduced by 10-50% among vaccinated elderly and comorbid individuals.

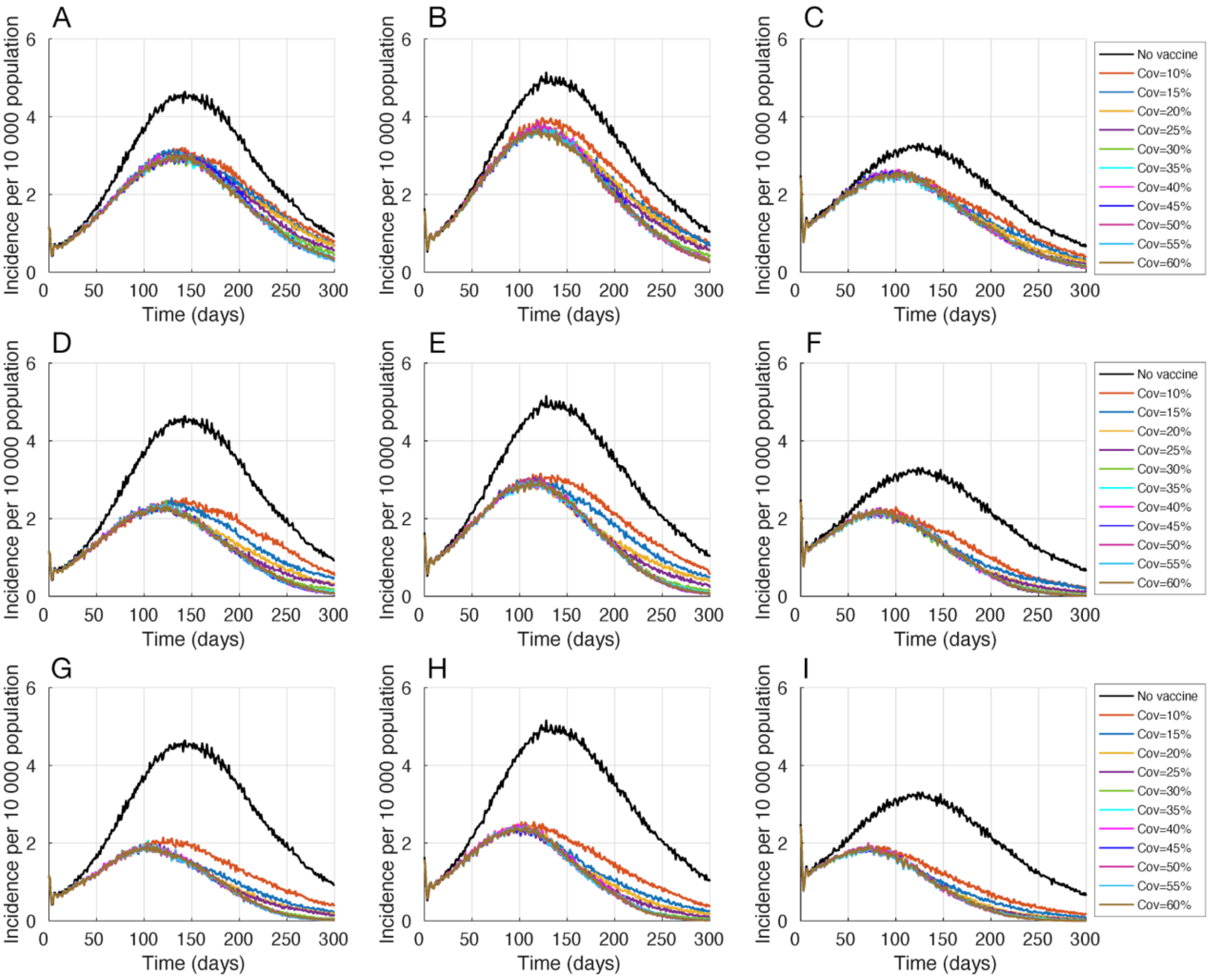


**Figure A1.** Projected daily incidence of COVID-19 per 10,000 population with 5% (A,D,G), 10% (B,E,H), and 20% (C,F,I) pre-existing immunity and different vaccination coverage in the range 10-60%. The protection efficacy of vaccines against infection is 0% (A,B,C), 50% lower than the efficacy against disease (D,E,F), and the same as efficacy against disease (G,H,I). Vaccine efficacy was reduced by 10-50% among vaccinated elderly and comorbid individuals.

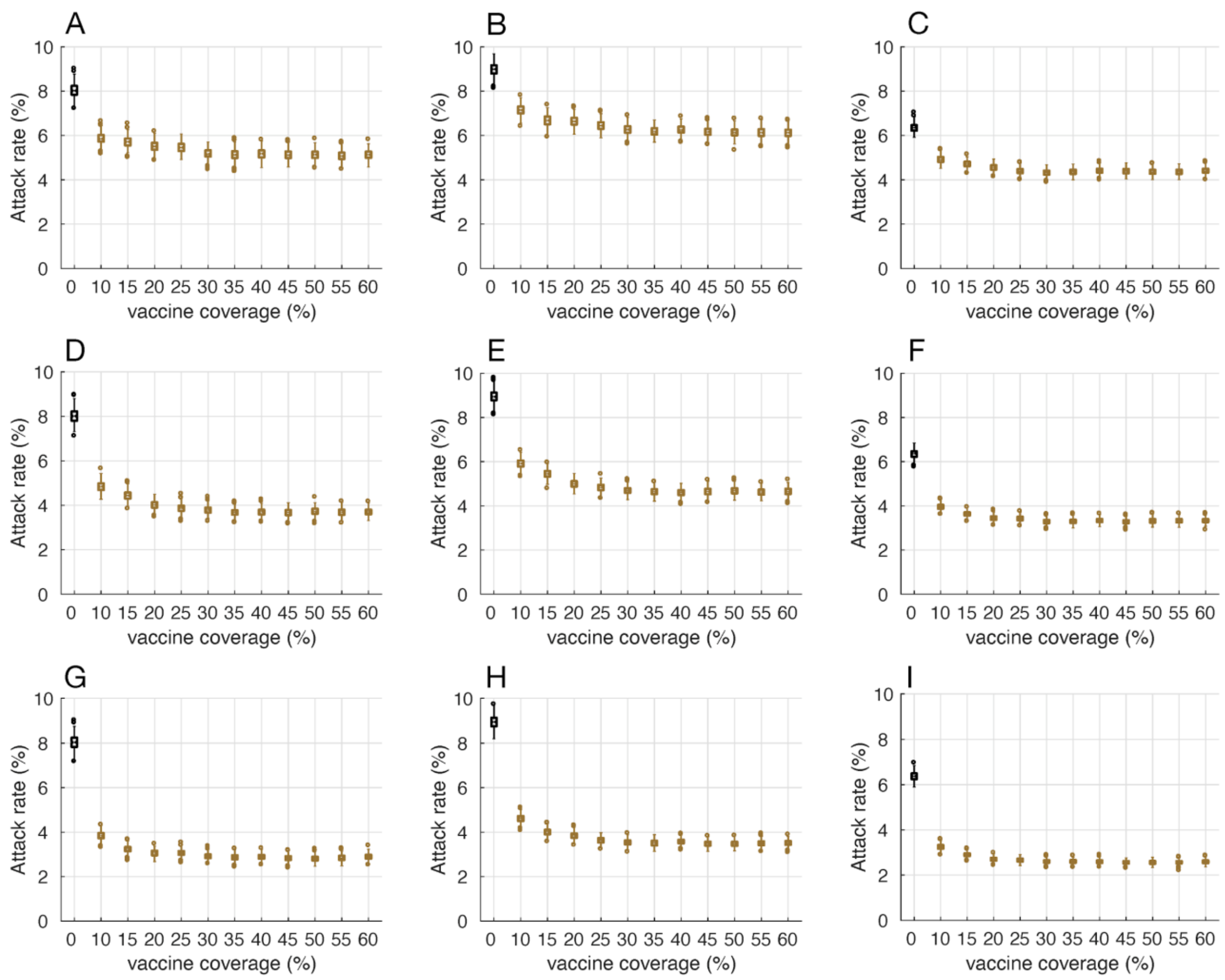


**Figure A2.** Projected attack rates with 5% (A,D,G), 10% (B,E,H), and 20% (C,F,I) pre-existing immunity and different vaccination coverage in the range 10-60%. The protection efficacy of vaccines against infection is 0% (A,B,C), 50% lower than the efficacy against disease (D,E,F), and the same as efficacy against disease (G,H,I). Vaccine efficacy was reduced by 10-50% among vaccinated elderly and comorbid individuals.

**Part 2:** There was no reduction of vaccine efficacy among vaccinated elderly and comorbid individuals.

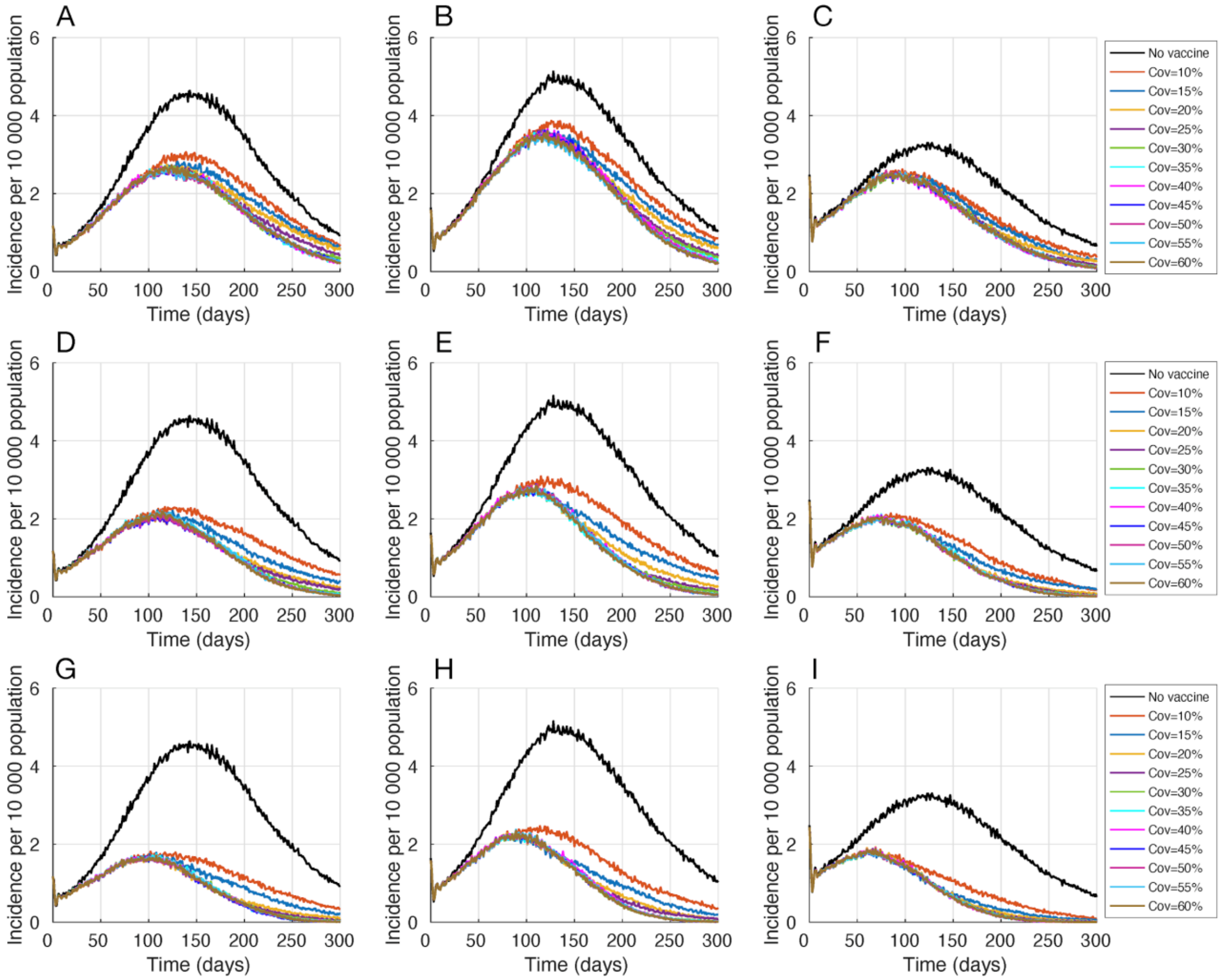


**Figure A3.** Projected daily incidence of COVID-19 per 10,000 population with 5% (A,D,G), 10% (B,E,H), and 20% (C,F,I) pre-existing immunity and different vaccination coverage in the range 10-60%. The protection efficacy of vaccines against infection is 0% (A,B,C), 50% lower than the efficacy against disease (D,E,F), and the same as efficacy against disease (G,H,I). There was no reduction of vaccine efficacy among vaccinated elderly and comorbid individuals.

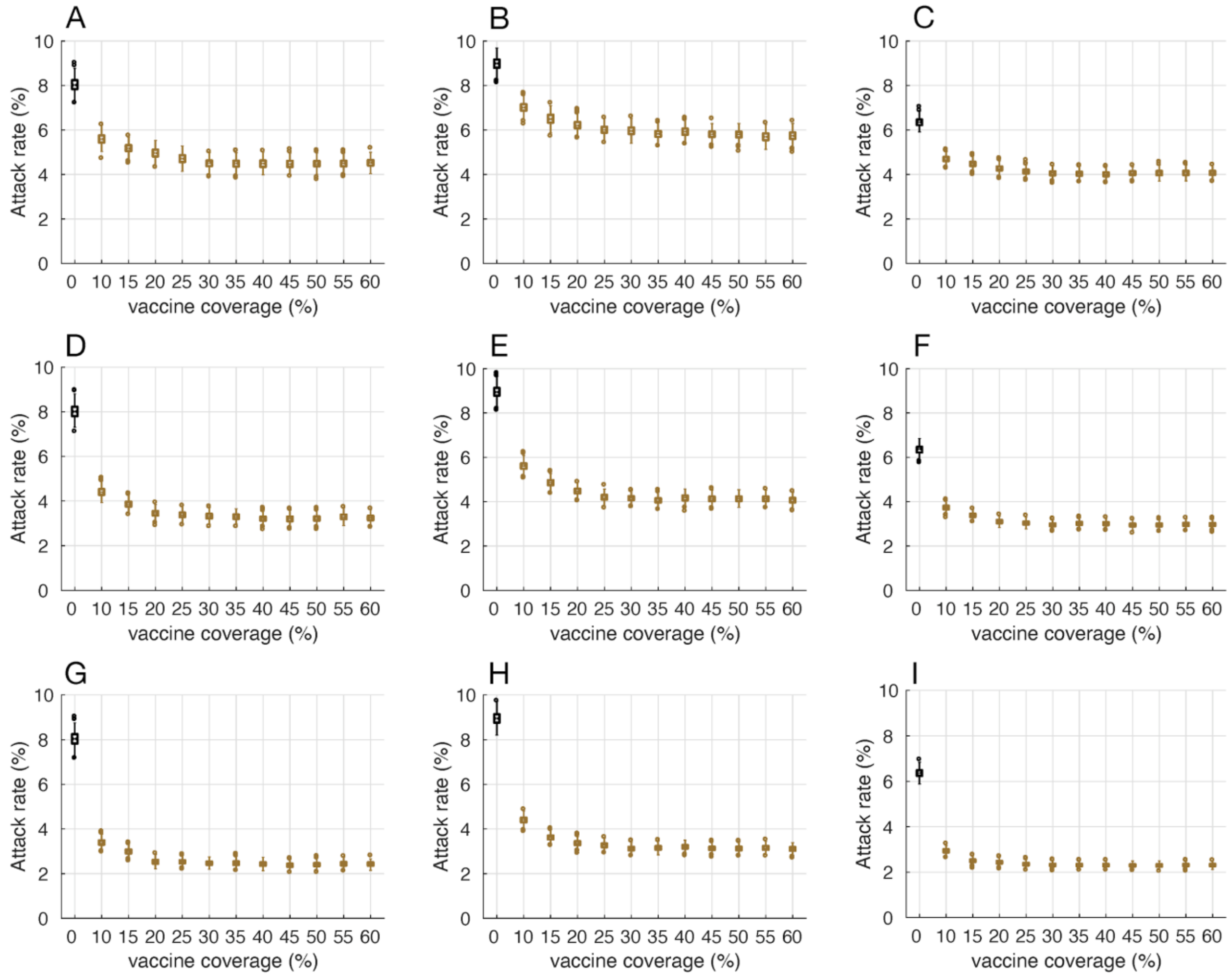


**Figure A4.** Projected attack rates with 5% (A,D,G), 10% (B,E,H), and 20% (C,F,I) pre-existing immunity and different vaccination coverage in the range 10-60%. The protection efficacy of vaccines against infection is 0% (A,B,C), 50% lower than the efficacy against disease (D,E,F), and the same as efficacy against disease (G,H,I). There was no reduction of vaccine efficacy among vaccinated elderly and comorbid individuals.

**Part 3:** Non-ICU hospitalizations, ICU hospitalizations, and deaths

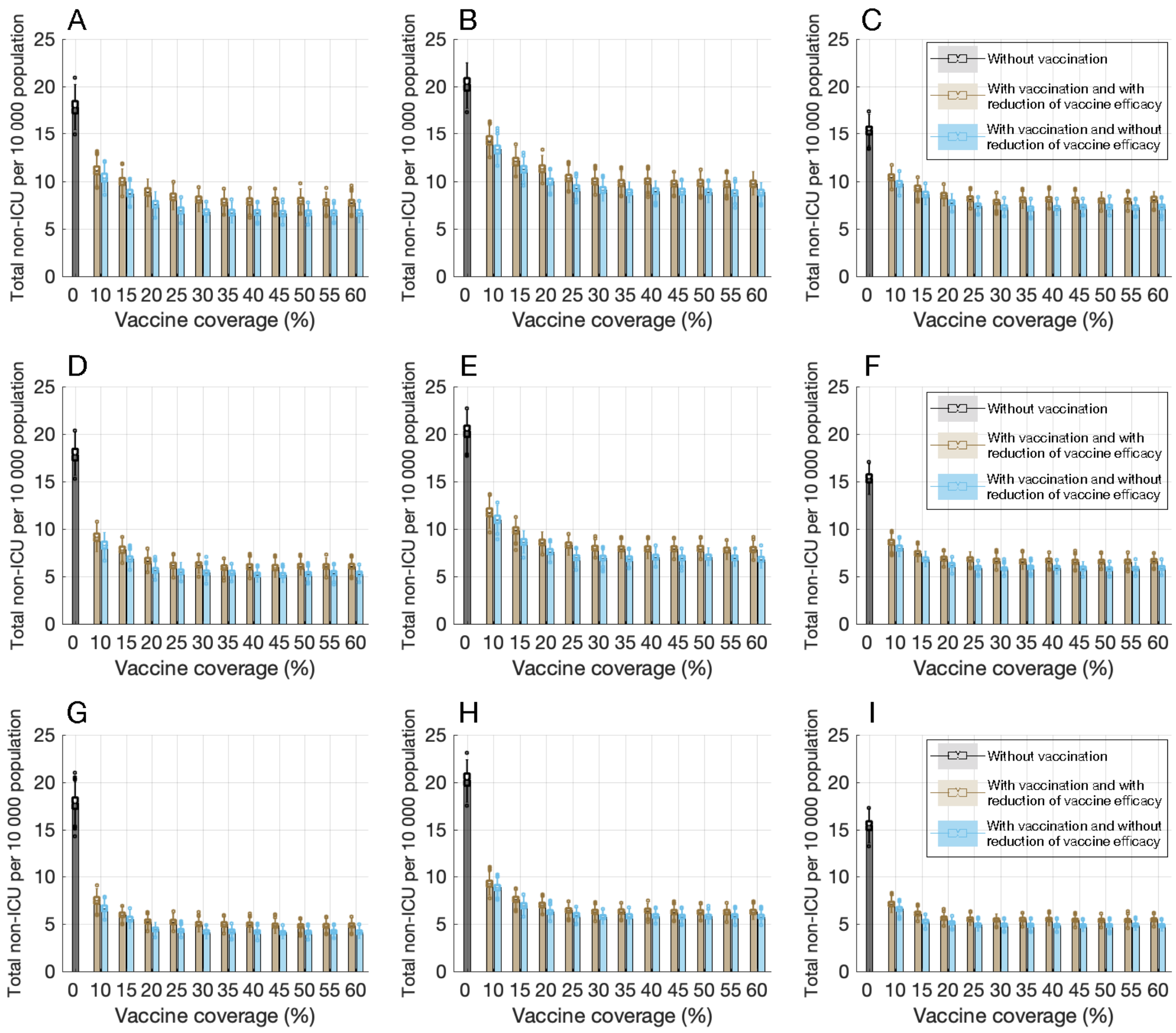


**Figure A5.** Projected total number of non-ICU hospitalizations per 10,000 populations with 5% (A,D,G), 10% (B,E,H), and 20% (C,F,I) pre-existing immunity and different vaccination coverage in the range 10-60% over 300 days. The protection efficacy of vaccines against infection is 0% (A,B,C), 50% lower than the efficacy against disease (D,E,F), and the same as efficacy against disease (G,H,I). Colored bars with vaccination correspond to scenarios with and without reduction of vaccine efficacy in comorbid individuals and the elderly.

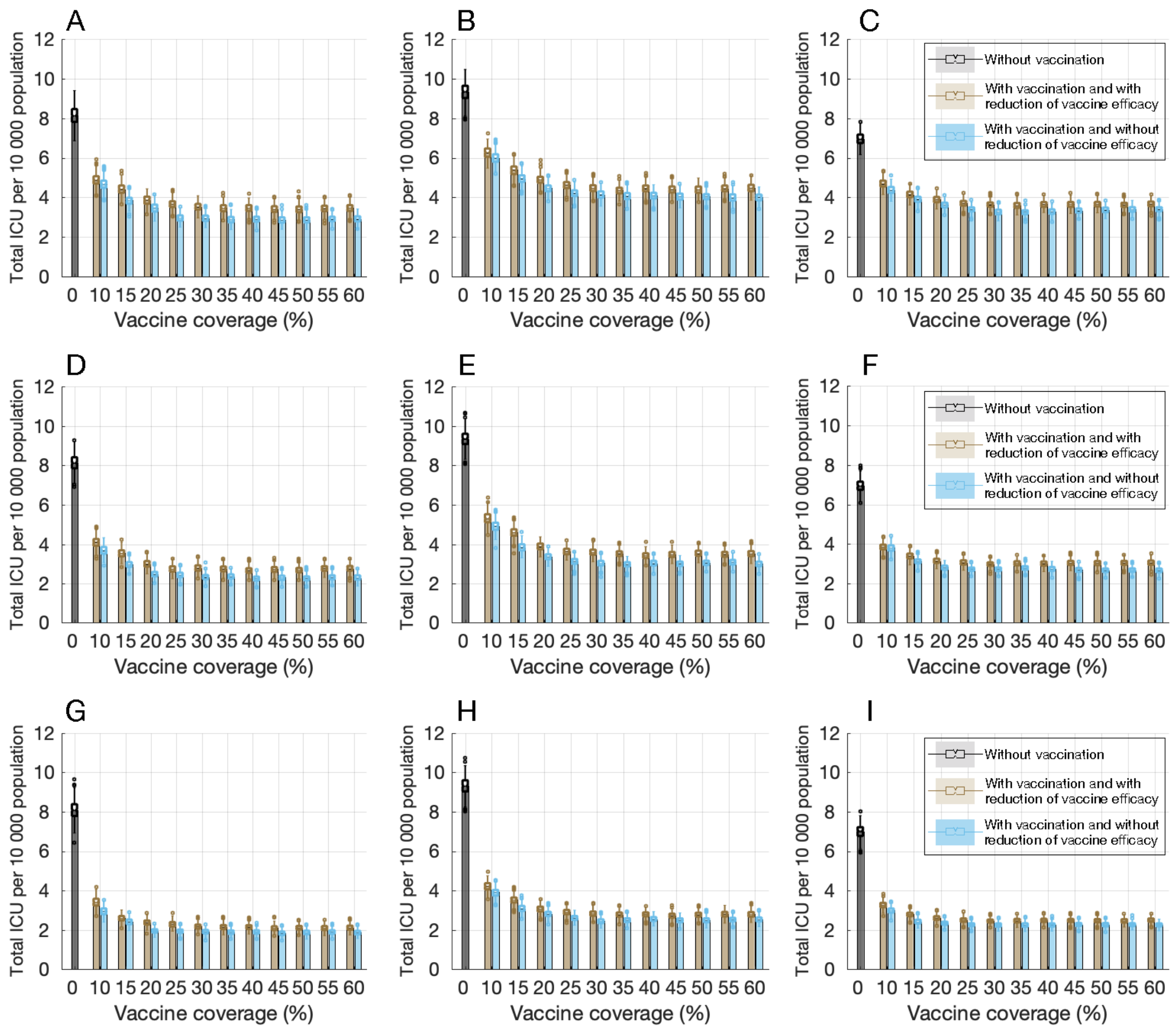


**Figure A6.** Projected total number of ICU hospitalizations per 10,000 populations with 5% (A,D,G), 10% (B,E,H), and 20% (C,F,I) pre-existing immunity and different vaccination coverage in the range 10-60% over 300 days. The protection efficacy of vaccines against infection is 0% (A,B,C), 50% lower than the efficacy against disease (D,E,F), and the same as efficacy against disease (G,H,I). Colored bars with vaccination correspond to scenarios with and without reduction of vaccine efficacy in comorbid individuals and the elderly.

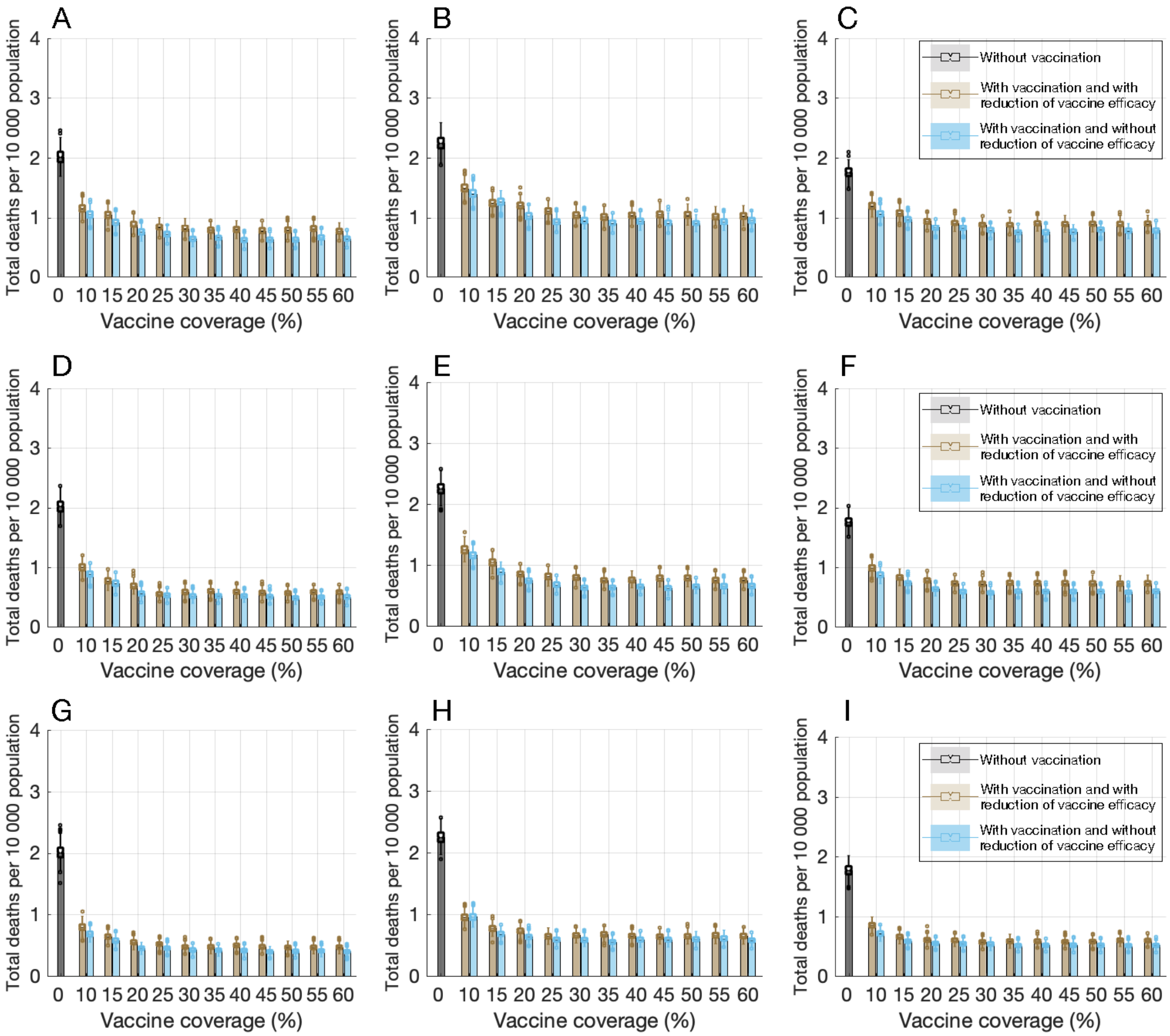


**Figure A7**. Projected total number of deaths per 10,000 populations with 5% (A,D,G), 10% (B,E,H), and 20% (C,F,I) pre-existing immunity and different vaccination coverage in the range 10-60% over 300 days. The protection efficacy of vaccines against infection is 0% (A,B,C), 50% lower than the efficacy against disease (D,E,F), and the same as efficacy against disease (G,H,I). Colored bars with vaccination correspond to scenarios with and without reduction of vaccine efficacy in comorbid individuals and the elderly.

### 5. Sensitivity analyses for vaccination scenarios with different coverages and protection efficacies against infection. Time interval between the two vaccine doses is 28 days.

**Part 1:** Vaccine efficacy was reduced by 10-50% among vaccinated elderly and comorbid individuals.

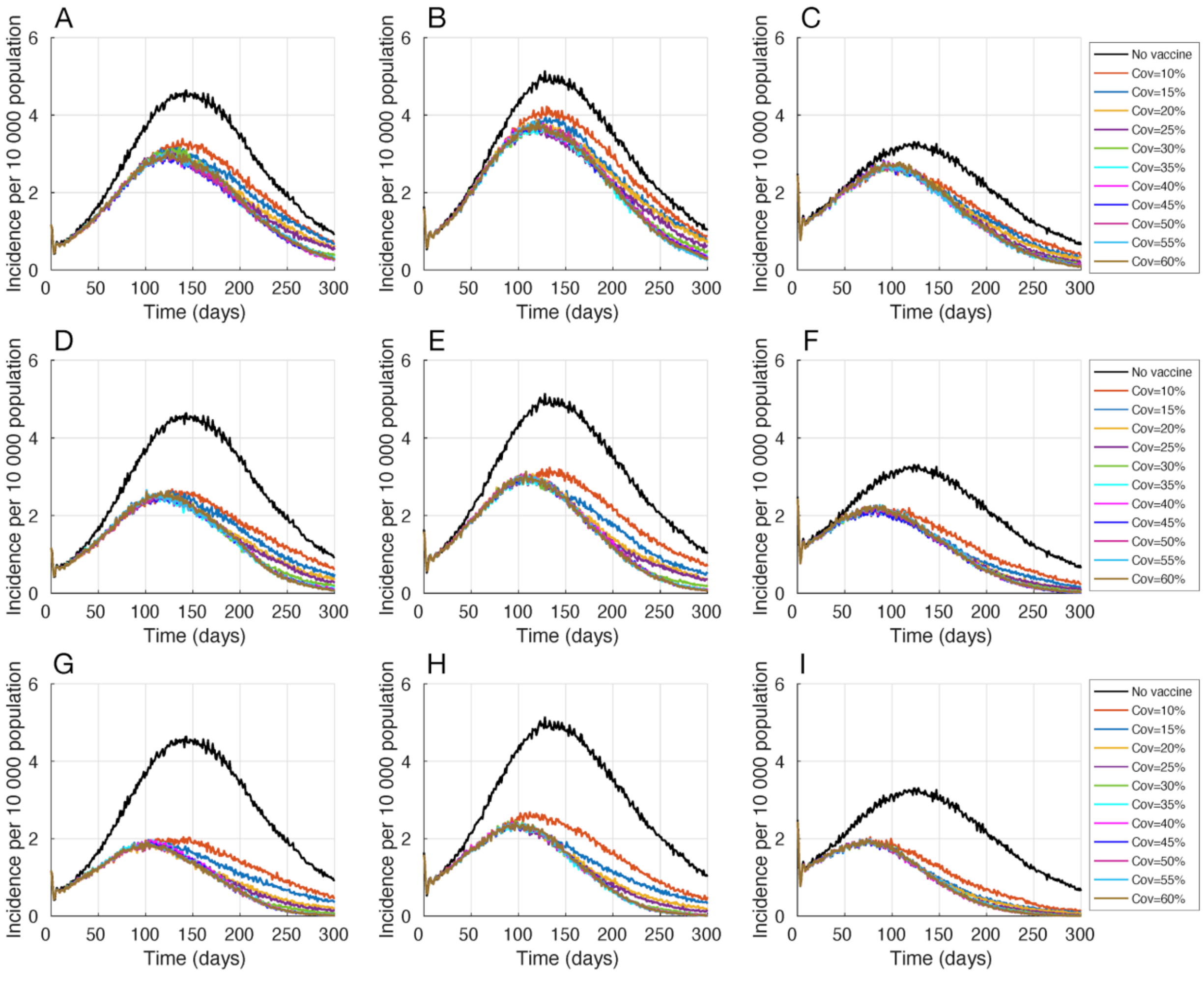


**Figure A8.** Projected daily incidence of COVID-19 per 10,000 population with 5% (A,D,G), 10% (B,E,H), and 20% (C,F,I) pre-existing immunity and different vaccination coverage in the range 10-60%. The protection efficacy of vaccines against infection is 0% (A,B,C), 50% lower than the efficacy against disease (D,E,F), and the same as efficacy against disease (G,H,I). Vaccine efficacy was reduced by 10-50% among vaccinated elderly and comorbid individuals.

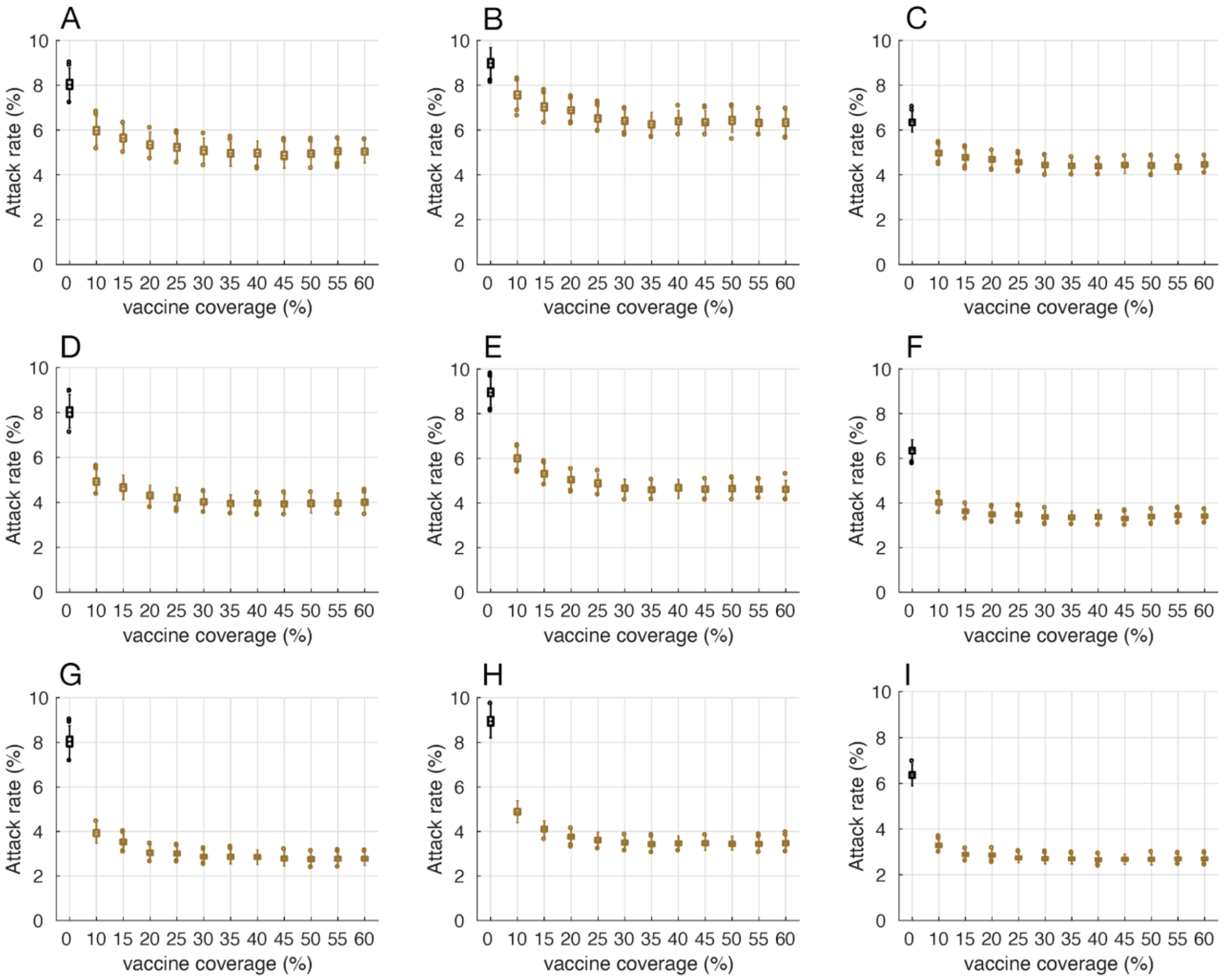


**Figure A9.** Projected attack rates with 5% (A,D,G), 10% (B,E,H), and 20% (C,F,I) pre-existing immunity and different vaccination coverage in the range 10-60%. The protection efficacy of vaccines against infection is 0% (A,B,C), 50% lower than the efficacy against disease (D,E,F), and the same as efficacy against disease (G,H,I). Vaccine efficacy was reduced by 10-50% among vaccinated elderly and comorbid individuals.

**Part 2:** There was no reduction of vaccine efficacy among vaccinated elderly and comorbid individuals.

**Figure A10.** Projected daily incid

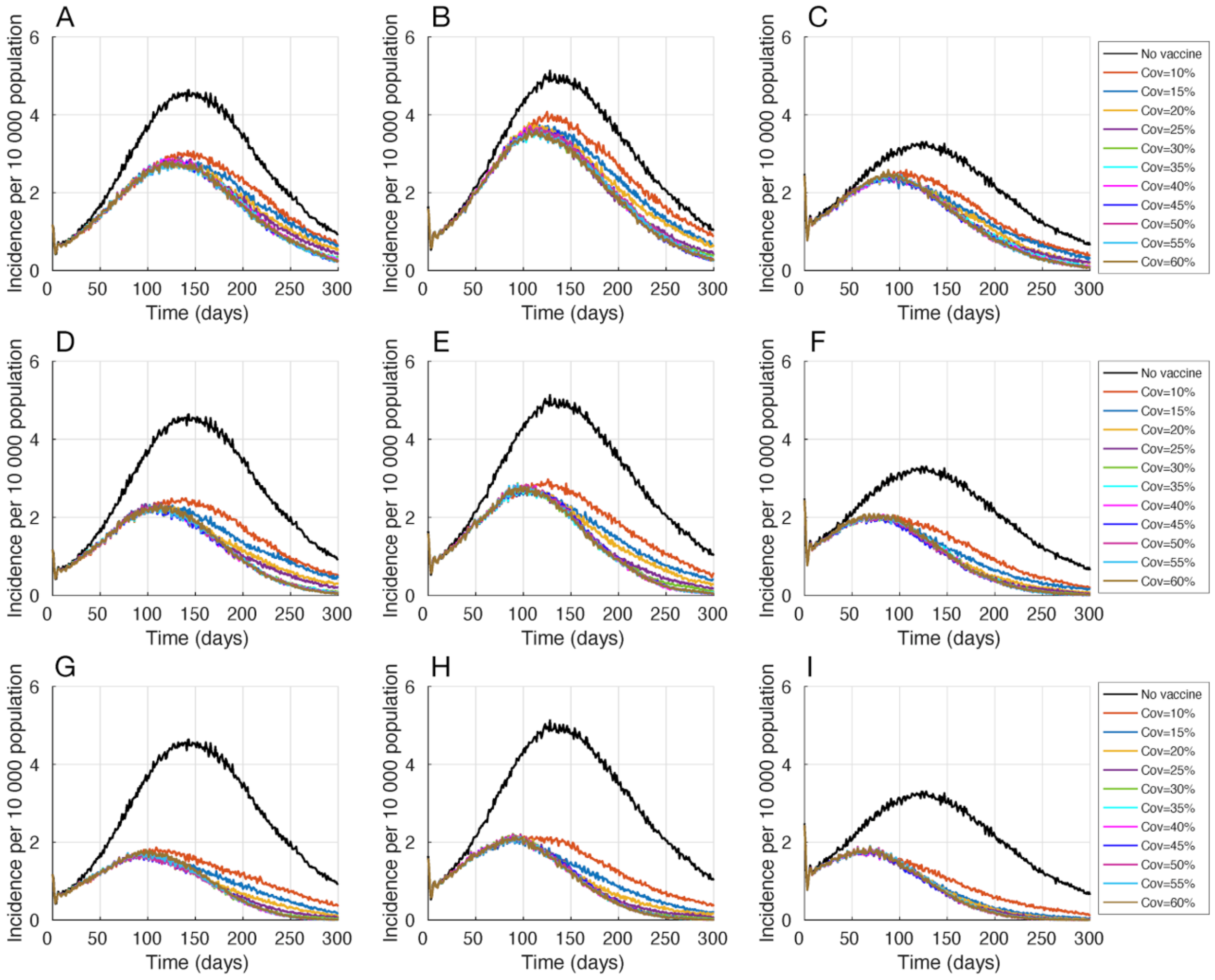


ence of COVID-19 per 10,000 population with 5% (A,D,G), 10% (B,E,H), and 20% (C,F,I) pre-existing immunity and different vaccination coverage in the range 10-60%. The protection efficacy of vaccines against infection is 0% (A,B,C), 50% lower than the efficacy against disease (D,E,F), and the same as efficacy against disease (G,H,I). There was no reduction of vaccine efficacy among vaccinated elderly and comorbid individuals.

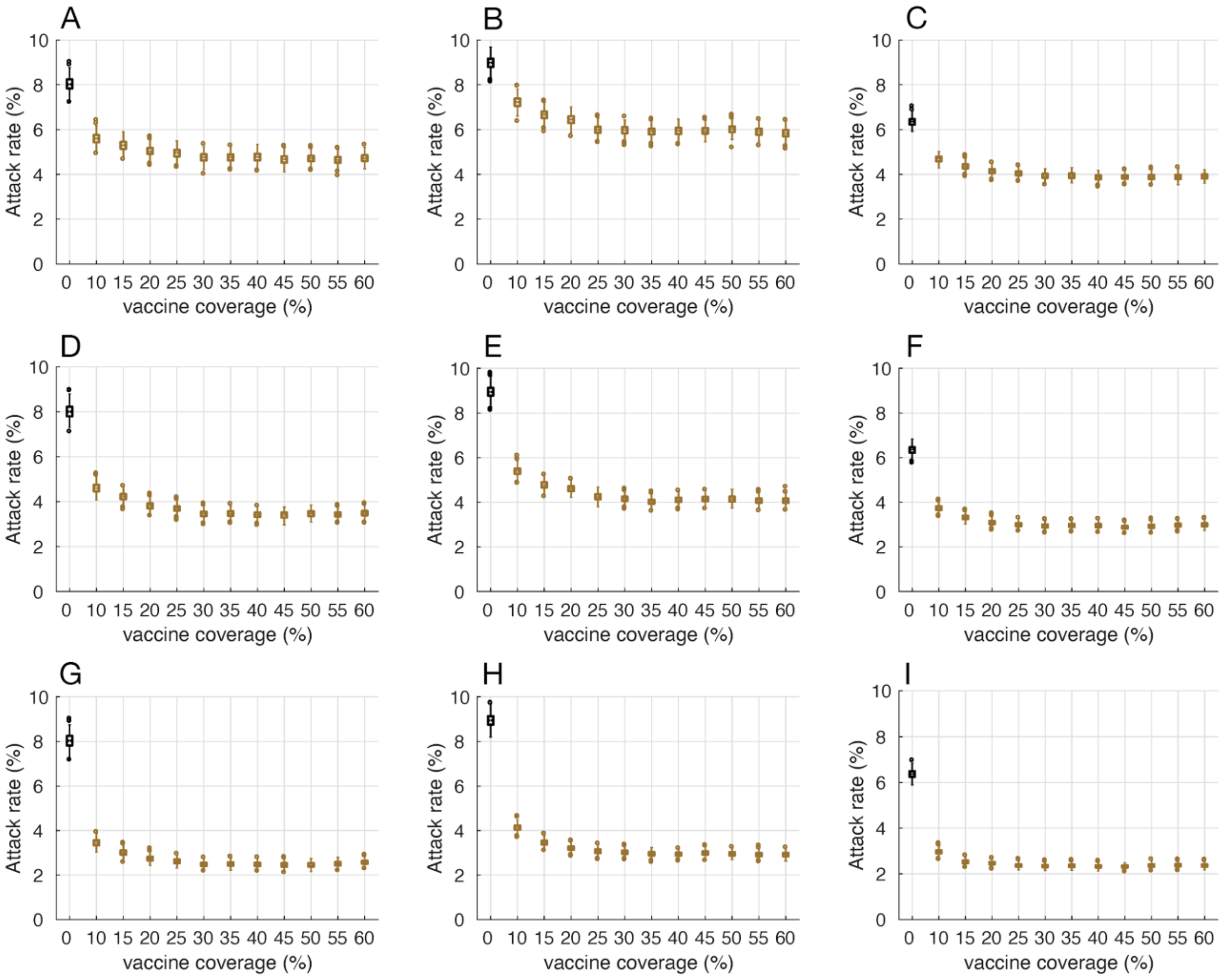


**Figure A11.** Projected attack rates with 5% (A,D,G), 10% (B,E,H), and 20% (C,F,I) pre-existing immunity and different vaccination coverage in the range 10-60%. The protection efficacy of vaccines against infection is 0% (A,B,C), 50% lower than the efficacy against disease (D,E,F), and the same as efficacy against disease (G,H,I). There was no reduction of vaccine efficacy among vaccinated elderly and comorbid individuals.

**Part 3:** Non-ICU hospitalizations, ICU hospitalizations, and deaths

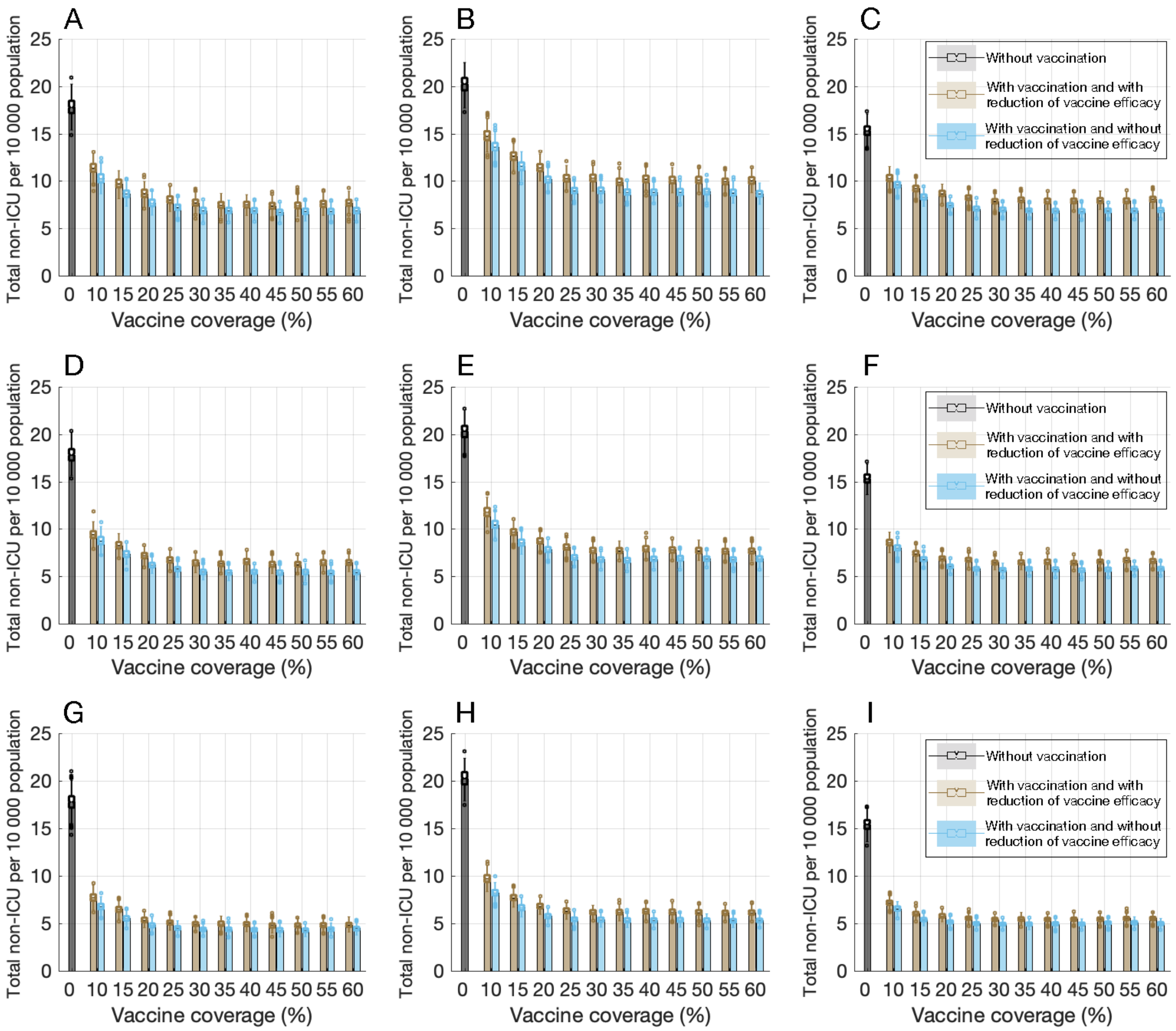


**Figure A12.** Projected total number of non-ICU hospitalizations per 10,000 populations with 5% (A,D,G), 10% (B,E,H), and 20% (C,F,I) pre-existing immunity and different vaccination coverage in the range 10-60% over 300 days. The protection efficacy of vaccines against infection is 0% (A,B,C), 50% lower than the efficacy against disease (D,E,F), and the same as efficacy against disease (G,H,I). Colored bars with vaccination correspond to scenarios with and without reduction of vaccine efficacy in comorbid individuals and the elderly.

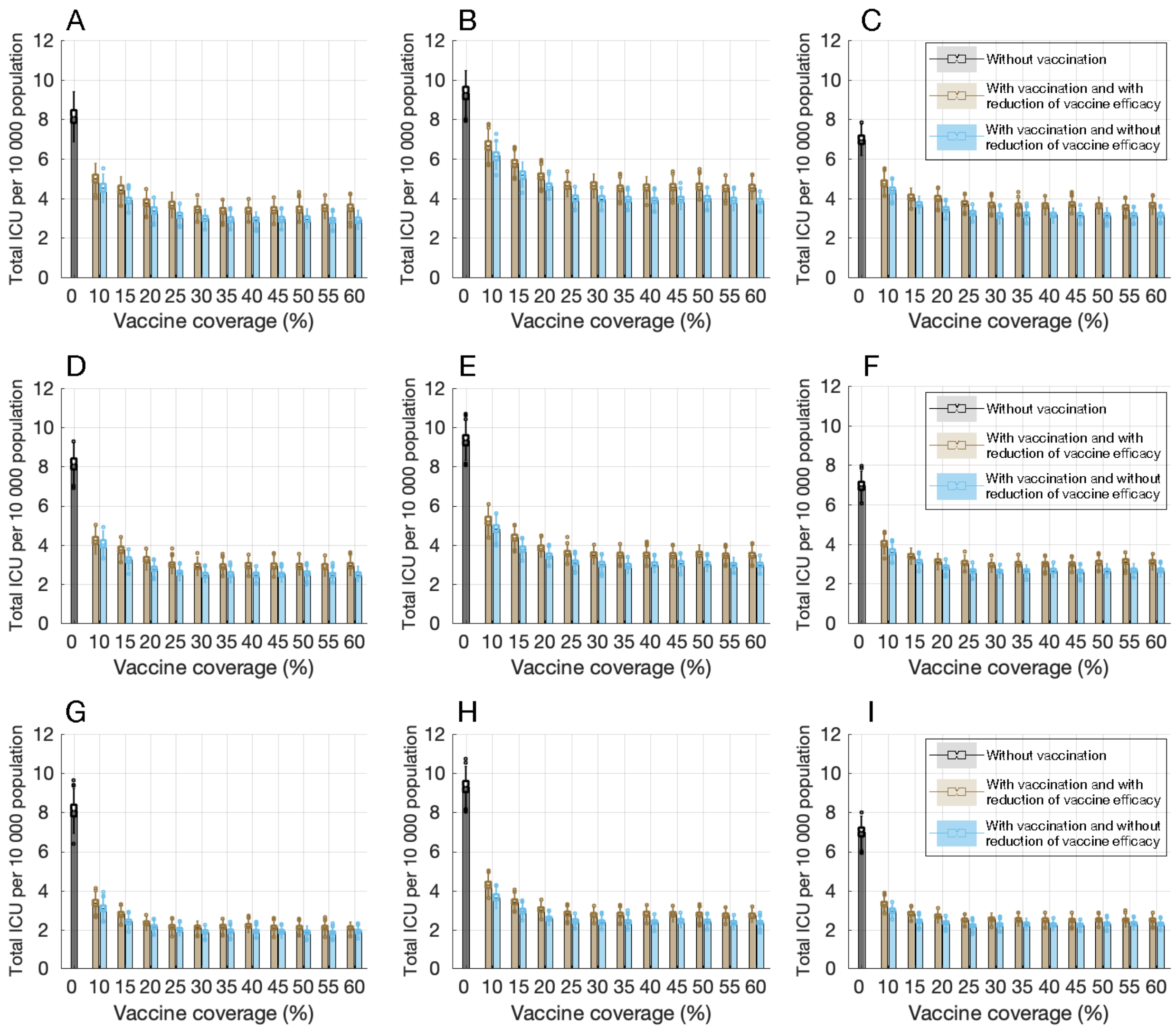


**Figure A13.** Projected total number of ICU hospitalizations per 10,000 populations with 5% (A,D,G), 10% (B,E,H), and 20% (C,F,I) pre-existing immunity and different vaccination coverage in the range 10-60% over 300 days. The protection efficacy of vaccines against infection is 0% (A,B,C), 50% lower than the efficacy against disease (D,E,F), and the same as efficacy against disease (G,H,I). Colored bars with vaccination correspond to scenarios with and without reduction of vaccine efficacy in comorbid individuals and the elderly.

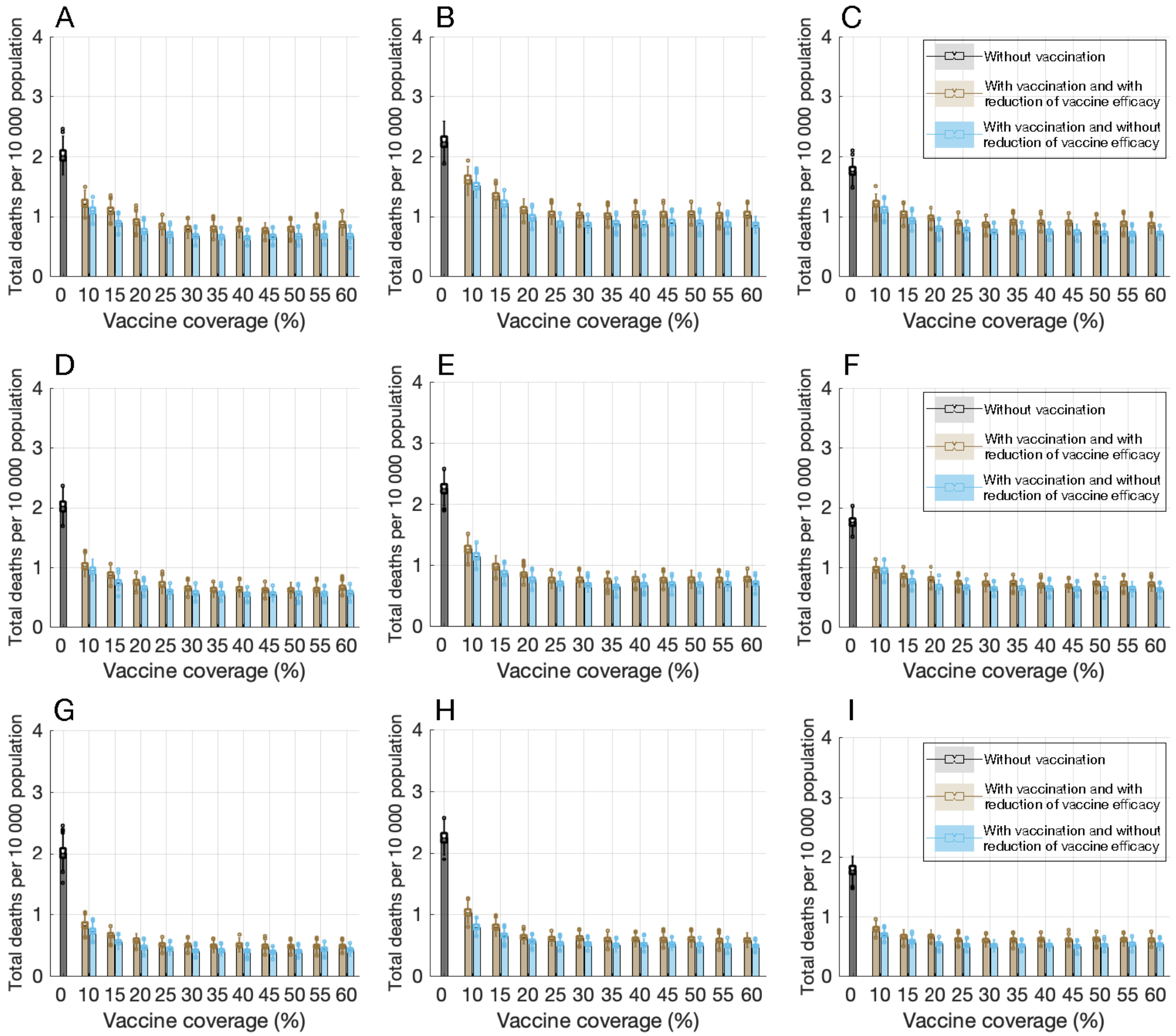


**Figure 14.** Projected total number of deaths per 10,000 populations with 5% (A,D,G), 10% (B,E,H), and 20% (C,F,I) pre-existing immunity and different vaccination coverage in the range 10-60% over 300 days. The protection efficacy of vaccines against infection is 0% (A,B,C), 50% lower than the efficacy against disease (D,E,F), and the same as efficacy against disease (G,H,I). Colored bars with vaccination correspond to scenarios with and without reduction of vaccine efficacy in comorbid individuals and the elderly.
